# Coping with Conflict: Short-Term Anxiolytic Medication Use Amidst National Stress in Israel

**DOI:** 10.1101/2024.10.04.24314902

**Authors:** Tal Patalon, Yaki Saciuk, Yogev Yonatan, Moshe Hoshen, Daniel Trotzky, Gal Pachys, Tsvi Fischel, Dorit Nitzan, Sivan Gazit

## Abstract

**Background:** Exposures to stress and traumatic events plays a significant role in triggering or precipitating anxiety. Nonetheless, these are often examined at the individual level, while societal-environmental exposures and their association with anxiety disorders are under-researched, especially in the Israeli context. This study leverages 19 years of longitudinal data from a large healthcare organization to examine the impact of national security instability on short-term anxiolytic purchases in Israel.

**Methods:** We conducted a retrospective cohort study using electronic medical records of over 1.1 million individuals from 2006 to 2024, examining rates of first-time and renewed use of anxiolytic medications of the benzodiazepines group during periods of armed conflict, including military operations and wars. Cox proportional hazards models were used to assess associations, adjusting for confounders such as age, sex, socioeconomic status, socioreligious sector, residence and previous psychiatric treatment.

**Results:** The risk for first purchase of an anxiety-relief short terms medication during military operations was 28% higher (HR 1.28, 95% CI: 1.21-1.34) compared to periods of relative national stability, after adjustments, and 44% higher during the Second Lebanon War (HR 1.44, 95% CI 1.27-1.62). The events of October 7^th^ were the most significant armed conflict increasing the risk for anxiety-related reaction necessitating treatment throughout the 19-years follow-up, with individuals at 317% increased risk for treatment initiation compared to periods of relative national stability (HR 4.17, 95% CI 3.97-4.38). Alongside a baseline increased risk for initiating anti-anxiety treatment, women experienced an additional elevated risk for anxiolytic therapy during times of national security threats, with 26% additional increased risk during military operations and an 81% increased risk following the events of October 7th. Residents of northern Israel had an increased risk of purchasing anxiolytics during the Second Lebanon War (HR 1.39, 95% CI: 1.12-1.72), while during military operations it was the residents of southern Israel who faced an increased risk for anxiolytic usage, with an HR of 1.18 (95% CI: 1.05-1.33). Conversely, the residential region did not significantly influence anti-anxiety treatment patterns following October 7th among residents of southern or northern Israel, compared to individuals living in central Israel, indicating a broader national impact beyond regional differences.

**Conclusions:** National armed conflicts significantly influence anxiolytic medication use in Israel, with the October 7th war showing the most pronounced effect. These findings highlight the need for comprehensive mental health interventions during times of national crisis, focusing on both short-term relief and long-term mental health support to prevent dependency and improve mental health outcomes in the wake of national crises.

## Introduction

Mental health challenges are complex, biopsychosocial conditions, acknowledged today as one of the most prevalent medical conditions. Of these, anxiety related disorders are some of the most common conditions, with a continuously rising prevalence.^1^ Acute symptoms of anxiety vary, from disturbing or intruding thoughts and emotions to psychophysiological manifestations, such as palpitations, gastrointestinal symptoms, excess sweating, lightheadedness and more. Long-term effects of anxiety have been linked to a plethora of chronic conditions, such as cardiovascular disorders^2–6^ neurodegenerative disorders including cognitive decline and dementia,^7,8^ as well as to other mental health conditions, such as depression.^9^

Exposures to stress and the experience of traumatic events have a significant role as triggers or precipitators of anxiety.^10,11^ Nonetheless, these are often examined at the individual level, leading to conjectures of psychoanalytical associations between exposure to stress, its idiosyncratic interpretations and the onset of anxiety.^12,13^ Contrarily, societal-environmental exposures and their association with anxiety disorders are under-researched, especially in the Israeli context.

Studies conducted with direct and indirect victims of national terror attacks (i.e., survivors, witnesses and close relatives of those who were killed) confirm that these individuals are at high risk of suffering from a distress-related reaction. In severe cases, such a distress reaction can result in varying severity levels of posttraumatic stress disorder (PTSD).^14^ Due to its prolonged political and security instability, Schechory-Biton et al have cited Israel as a “stress laboratory” for the study of war- and terror-related stress, as acts of terrorism and national insecurity are part of daily life.^15^ Correspondingly, Ayer et al conducted a systematic review of the psychological consequences of the Israeli-Palestinian conflict, including over fifty studies, of which more than 20 focused on PTSD. The authors concluded that there was a positive correlation between exposure to regional conflicts and detrimental effects on psychological wellbeing beyond the diagnosed psychiatric disorders. However, the absence of longitudinal data and lack of an effect size, limited the findings of the study.^16^

Given the paucity of real-world studies leveraging longitudinal data to examine exposure to stress at the societal-national (rather than the individual psychological one) level, and resulting anxiety, we conducted this retrospective analysis. Our aim was to explore the association between periods of peaked national stress in the past 19 years, including the latest events of October 7^th^, and the incidence of anxiety reactions necessitating medical intervention with short-term anxiolytics.

## Methods

### Data sources

Maccabi Healthcare Services (MHS) is the second largest Israeli health fund (insurer-provider), covering 26.7% of the Israeli population through a nationwide network. It has maintained a centralized database of electronic medical records (EMRs) for over thirty years, allowing a substantial longitudinal follow-up on a stable population, with less than 1% yearly disengagement rate. The database holds extensive information, automatically captured, including demographic data, diagnoses and procedures, imaging and laboratory tests, as well as dispensed medications.

### Data extraction

Anonymized EMRs were retrieved from MHS from January 1, 2006 through July 31^st^, 2024. Individual-level data included demographic information, explicitly sex, year of birth, social-religious sector (ultra-orthodox Jew, Arab, and general population), geographical district of residence and socioeconomic status (SES). SES is assigned by the Israel’s Central Bureau of Statistics,^17^ and measured from 1 (lowest) to 10, where index is calculated based on several parameters, including household income, educational qualifications, household crowding and car ownership. Data also included medication purchases, where these were coded by the Anatomical Therapeutic Chemical (ATC) medications codes. Data also included a history of using psychiatric medication.

### Study population

Study population included all MHS members aged 21 years or older by January 2006. Participants were excluded from the study if they joined MHS after the age of 25, in order to allow follow-up from young adulthood and thus to allow for sufficient history collection that will differentiate newly purchased medications from chronic ones.

Individuals were censored in cases of disengagement with the health fund or death. All sectors, socioeconomic statuses and both genders of the population were included.

### Study design

We conducted a retrospective cohort study, examining rates of anxiolytic medication purchases by MHS members between 2006 and 2024. Individuals were examined across different time periods, where periods of armed conflict were flagged as such by their calendar date, focusing on the first day of events and up to 3 weeks since, marking a change from baseline. That is, an event was defined by the week it started through the following two weeks. We differentiated between large military operations and war, given their often different duration and different levels of direct security effect on civilian population. The former category included four military operations: Cast Lead (2008, in Hebrew “Oferet Yetsuka”),^18^ Pillars of Defense (2012, “Amud Anan”),^19^ Protective Edge (2014, “Tsuk Eitan”) ^20^ and Guardian of the Walls (2021, “Shomer Homot”).^21^ Two wars were included in the analysis, The Second Lebanon War (2006)^22^ and the current Israel-Hamas War (2023, “Haravot Barzel” or “October 7^th^ war”).^23^ Additionally, given the overwhelming reaction in Israel to recent events in both military and civilian aspects, we further differentiated between the Second Lebanon War, and the events of October 7^th^, thus creating two distinct war sub-categories.

### Measured outcome

Our study examined the association between the exposure variables – ‘military operations’, ‘second Lebanon war’ and ‘October 7^th^ war’ – and the incidence rates of an acute anxiety-related or stress-related reaction necessitating pharmacological intervention with short-term anxiety-related medications of the benzodiazepine pharmacological group. We further differentiated between a first-ever purchase and a repeated purchase after a one-year washout period (of zero purchases).

### Statistical analysis

#### Primary analysis

We first examined the unadjusted incidence rates (IRs) of anxiolytics purchases per 100,000 person days at risk to demonstrate first-time purchases over time. Next, an extended Cox proportional hazards model was fit to the data, adjusting for possible confounders, including age, biological sex, socio-religious sector, socioeconomic status, general area of residence, residence within the Gaza envelope and usage of psychiatric medications at any time up and including the exposure weeks, testing for time to onset of a first anxiolytic purchase. The near-Gaza area (Gaza envelope), was designated separately, this area of residence within 7 kilometers of the Gaza Strip, has been facing more frequent armed attacks than other civilian areas of residence in recent years. Ages were grouped into 21-24, 25-39, 40-59, and 60 or older to reflect the Israeli social context, where military service is mandatory. These categories account for different stages of military involvement: individuals aged 21-24 are often still in active duty (serving as officers or non-comissioned officers after completing mandatory service), those aged 25-39 are generally in reserve duty (which typically ends at 40), those aged 40-59 are often parents of children in active military service, and those aged 60 and older less directly impacted by military service.Lastly, we adjusted for calendar time using restricted cubic splines for the continuous time indicator to allow for nonlinear time trends.

We thus calculated the hazard ratio (HR), comparing periods of national conflict to corresponding relative national stability (between operations and wars), where the latter times were the reference group. We further included additional regressions with interaction terms between the above-mentioned covariates and the exposure events, namely times of conflict. Additionally, we conducted a stratified analysis by the different age groups, to further test their reaction to these national conflicts. Lastly, we stratified our analysis by socio-religious sector, examining the Arab and general Jewish population populations separately, given their presumed different behaviors as it comes to seeking pharmacological interventions for mental health challenges, as well as the former’s general non-participation in military service.

#### Secondary analysis

Our secondary analysis implemented the same design, while examining the different patterns of anxiolytic medication purchases: a repeated purchase (rather than an individual’s first purchase), that is a renewed usage of anxiolytic medications after at least one-year ‘washout’ period during which no such treatment was sought. All other variables were similar.

#### Supplementary analysis: negative controls

To bolster the validity of our associations, we employed a Negative Controls approaches,^24^ swapping the exposure with an event presumably unrelated to the measured outcome (often referred to as ‘exposure negative control’). The analysis is then performed while maintaining the consistency of all other factors in the model. If results demonstrate that previously observed associations disappear in this altered scenario, this strengthens the reliability of the inference. In this study, we used the shifting of national conflict dates by exactly one year (to preserve seasonality) as exposure negative controls, flagging them in the same manner to the primary and secondary analyses.

Analyses were conducted using Python version 3.10.9 with statsmodels package and R (4.3.0 using the survival package) and in accordance with the STROBE statement.

### Ethics declaration

This study was approved by the MHS Institutional Review Board (IRB). Due to the retrospective design of the study, informed consent was waived by the IRB, and all identifying details of the participants were removed before analysis.

### Data and Code availability statement

According to the Israel Ministry of Health regulations, individual-level data cannot be shared openly. Specific requests for remote access to de-identified community-level data or code used should be referred to Maccabi Healthcare Services Research and Innovation Center.

## Results

### Primary analysis

A total of 1,142,6441 individuals contributed 12,027,931 person-years at risk during the 19-year study period, with 9,973,849 person years were at risk of first purchase. 173,120 new purchases of anxiolytic medications occurred during these years, with an overall slight gradual since 2006 in absolute numbers. A marked spike in first-time and anxiolytic purchases can be seen in late 2023, with smaller peaks in 2014 and 2006 (**Figure 1**). The sharp increase circa October 2023 far exceeds all previous periods throughout the 19-years examined period. When particularly examining absolute purchases in the weeks pre- and post-armed conflicts on a weekly resolution (**Figure 2**), an evident temporal increase in first-time purchases is evident around the first weeks following October 7^th^ war (and later decreasing) as well as during Protective Edge.

**Figure 1.**
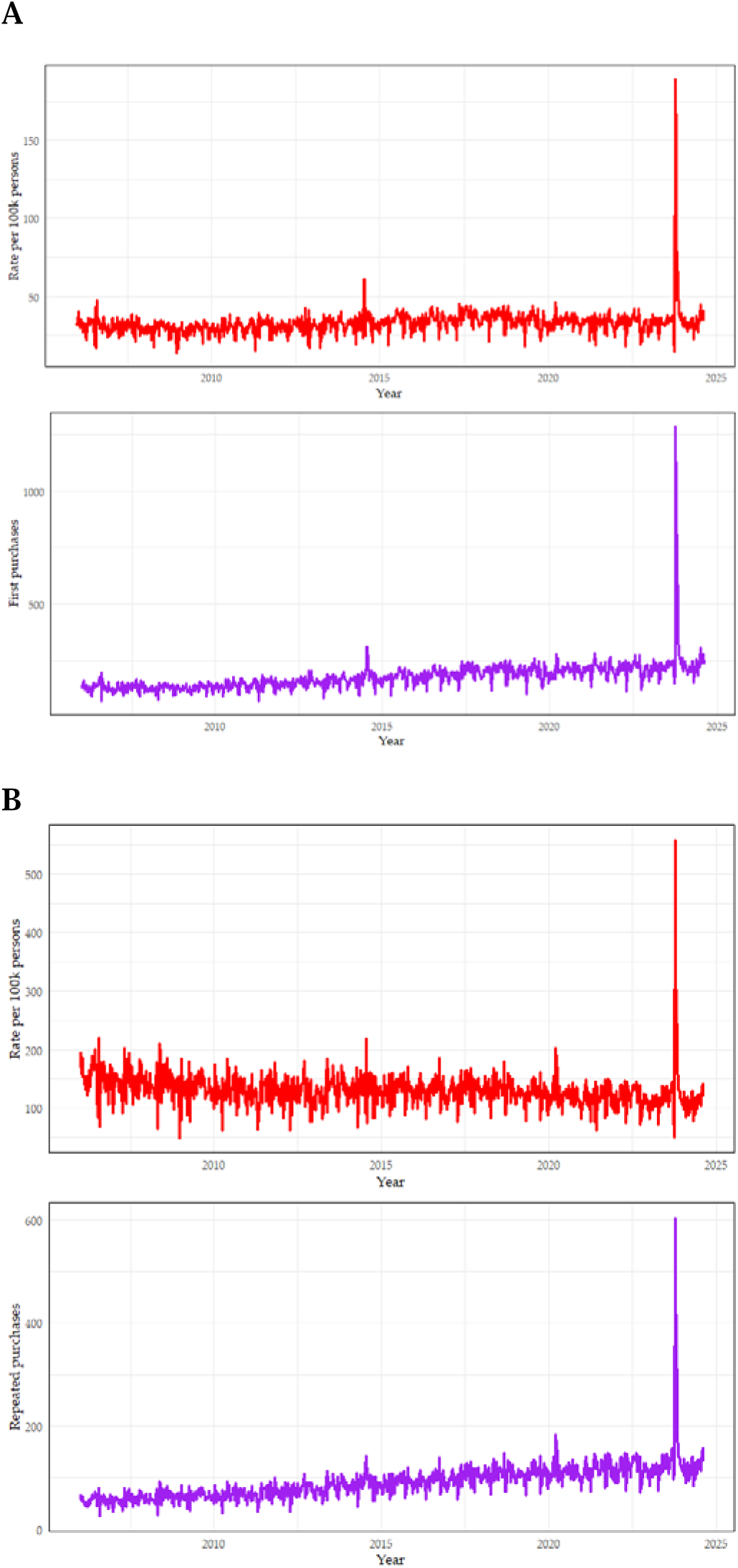
First purchases (A panels) and renewed purchases (B panels) of short-term anxiolytic medications by MHS members between 2006 and 2024, as unadjusted incidence rate per 100,000 individuals and in absolute numbers.

**Figure 2.**
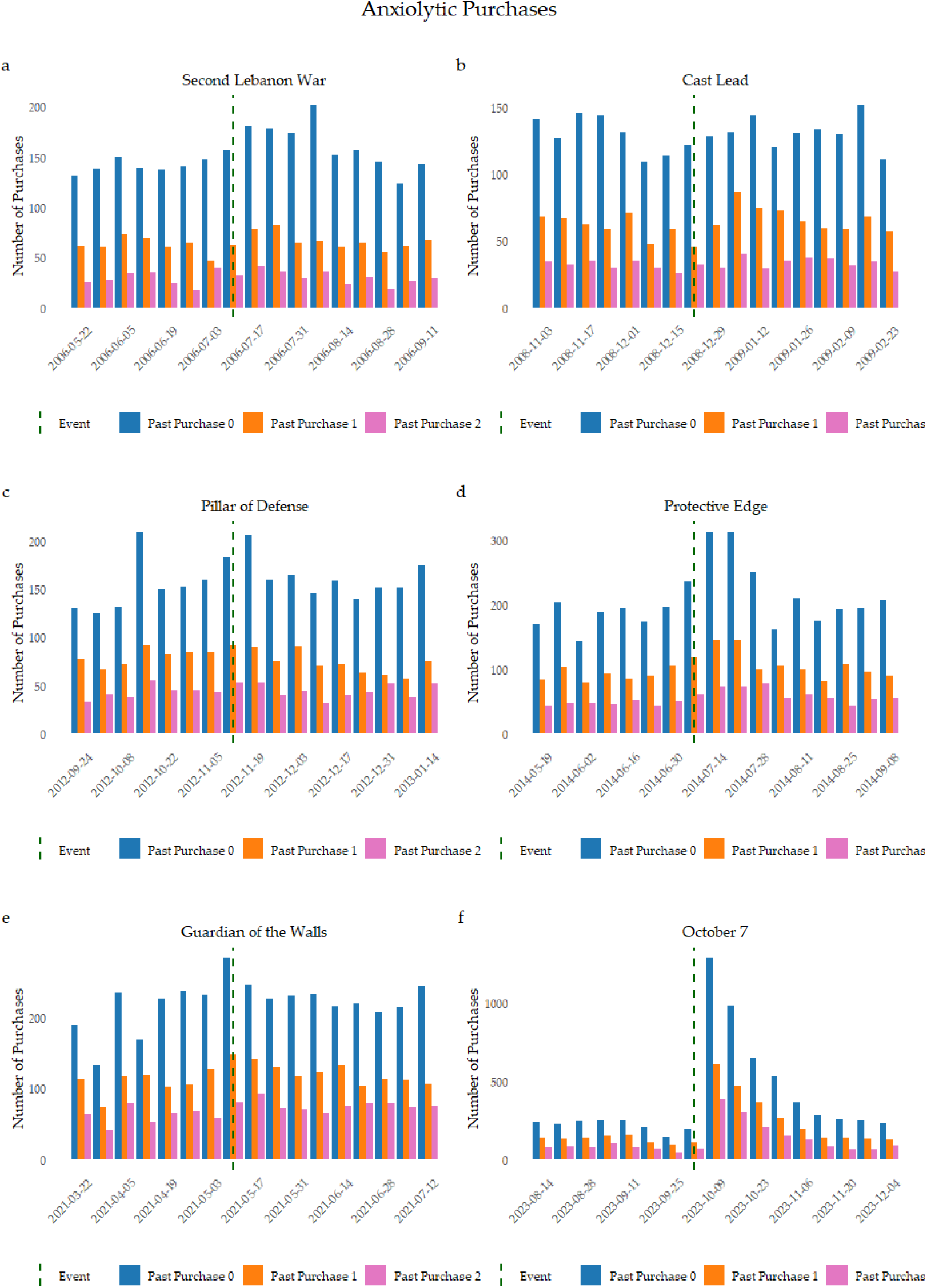
Absolute purchases of anxiolytics in the weeks prior to and after national armed conflicts in Israel, stratified by first purchase, repeated first purchase and repeated purchase after at least two previous purchases. Each repeated purchase was preceded by at least one year washout period.

The population characteristics of individuals with and without a purchase of anxiolytic at the first and last year of examined armed conflicts can be seen in **Table 1**, demonstrating that the mean age of first purchase of anxiolytics occurred in the late 30’s, with a strong female predominance, with fewer purchases in the Arab and Ultra-Orthodox Jewish populations.

**Table 1.** Characteristics of MHS participants in 2006 (year of first military conflict examined in this study) and 2023 (last year in which a military conflicted broke), with and without a purchase of anxiolytic medication. Data represent number (%) of participants, unless stated otherwise.

| 2006 |  |  |  |
| --- | --- | --- | --- |
|  | Didn't Purchase<br>(N=432442) | Purchased<br>(N=7072) | Overall<br>(N=439514) |
| **Age** |  |  |  |
| Mean (SD) | 32.2 (8.86) | 36.4 (11.2) | 32.3 (8.91) |
| Median [Min, Max] | 31.0 [21.0, 87.0] | 34.0 [21.0, 80.0] | 31.0 [21.0, 87.0] |
| **Sex** |  |  |  |
| M | 195537 (45.2%) | 2509 (35.5%) | 198046 (45.1%) |
| F | 236905 (54.8%) | 4563 (64.5%) | 241468 (54.9%) |
| **SES** |  |  |  |
| High (8-10) | 139180 (32.2%) | 2389 (33.8%) | 141569 (32.2%) |
| Medium (5-7) | 223329 (51.6%) | 3699 (52.3%) | 227028 (51.7%) |
| Low (1-4) | 69933 (16.2%) | 984 (13.9%) | 70917 (16.1%) |
| **Sector** |  |  |  |
| General | 377591 (87.3%) | 6306 (89.2%) | 383897 (87.3%) |
| Ultraorthodox Jew | 35847 (8.3%) | 496 (7.0%) | 36343 (8.3%) |
| Arab | 19004 (4.4%) | 270 (3.8%) | 19274 (4.4%) |
| 2023 |  |  |  |
|  | Didn't Purchase<br>(N=692837) | Purchased<br>(N=14383) | Overall<br>(N=707220) |
| **Age** |  |  |  |
| Mean (SD) | 35.3 (12.2) | 37.0 (12.6) | 35.4 (12.2) |
| Median [Min, Max] | 32.0 [21.0, 95.0] | 34.0 [21.0, 91.0] | 32.0 [21.0, 95.0] |
| **Sex** |  |  |  |
| M | 336206 (48.5%) | 5280 (36.7%) | 341486 (48.3%) |
| F | 356631 (51.5%) | 9103 (63.3%) | 365734 (51.7%) |
| **SES** |  |  |  |
| High (8-10) | 237960 (34.3%) | 5715 (39.7%) | 243675 (34.5%) |
| Medium (5-7) | 310702 (44.8%) | 6303 (43.8%) | 317005 (44.8%) |
| Low (1-4) | 144175 (20.8%) | 2365 (16.4%) | 146540 (20.7%) |
| **Sector** |  |  |  |
| General | 576000 (83.1%) | 12762 (88.7%) | 588762 (83.3%) |
| Ultraorthodox Jew | 69517 (10.0%) | 1108 (7.7%) | 70625 (10.0%) |
| Arab | 47320 (6.8%) | 513 (3.6%) | 47833 (6.8%) |

<sup>1</sup>SD - standard deviation; <sup>2</sup>SES - socioeconomic status.

When fitting a Cox regression model to the data and adjusting for above-mentioned possible confounders, we found an overall increased risk for anti-anxiety medication with ages, where individuals 60 years or older were 2.02 (95% CI: 1.95-2.09) times more likely to be treated compared to 21–24-year-olds (**Table 2**, **Figure 3**).

**Figure 3.**
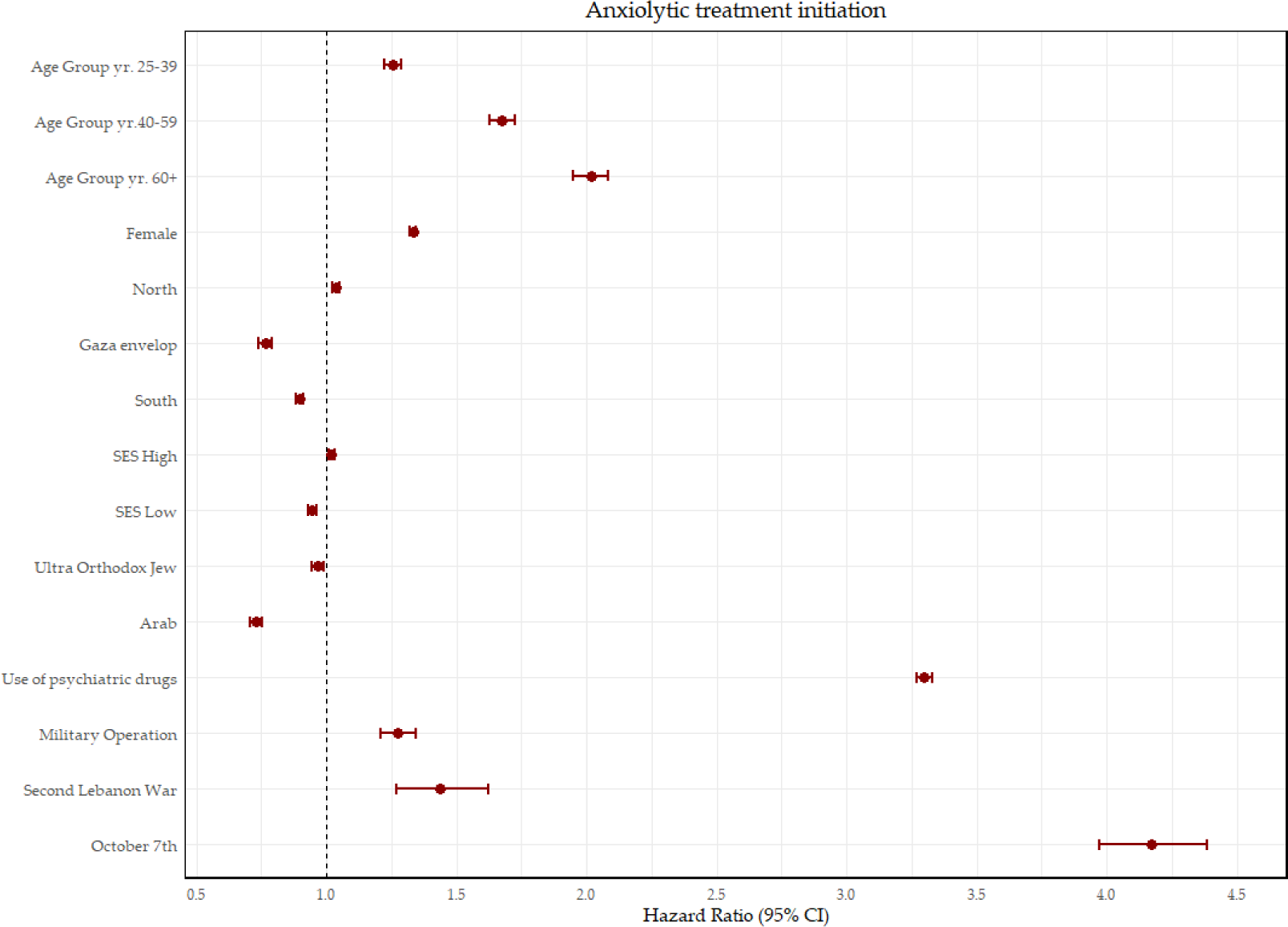
Forest Plot of HRs and 95% confidence intervals, primary analyses. SES - socioeconomic status.

**Table 2.** Cox regression analysis with adjusted Hazard Ratios (HRs) for first purchase of anxiolytic medications (primary analysis) between 2006-2024.

| Variable | Category | HR | P-value | 95% CI |
| --- | --- | --- | --- | --- |
| Age group, yr. | 25-39 | 1.26 | <0.001 | 1.22 - 1.29 |
|  | 40-59 | 1.68 | <0.001 | 1.63 - 1.72 |
|  | 60+ | 2.02 | <0.001 | 1.95 - 2.09 |
|  | 21-24 | Ref |  |  |
| Biological sex | Female | 1.33 | <0.001 | 1.32 - 1.35 |
|  | Male | Ref |  |  |
| District | North | 1.04 | <0.001 | 1.02 - 1.05 |
|  | South | 0.90 | <0.001 | 0.88 - 0.91 |
|  | Gaza envelope | 0.77 | <0.001 | 0.74 - 0.79 |
|  | Center | Ref |  |  |
| SES | Low | 0.95 | <0.001 | 0.93 - 0.96 |
|  | Medium | Ref |  |  |
|  | High | 1.02 | 0.001 | 1.01 - 1.03 |
| Socio-religious sector | Ultra-Orthodox Jew | 0.97 | <0.001 | 0.95 - 0.99 |
|  | Arab | 0.73 | 0.004 | 0.71 - 0.75 |
|  | General Jewish | Ref |  |  |
| Use of psychiatric drugs | Yes | 3.30 | <0.001 | 3.27 - 3.33 |
|  | No | Ref |  |  |
| Armed conflict | Second Lebanon War | 1.44 | <0.001 | 1.27 - 1.62 |
|  | Military operation | 1.28 | <0.001 | 1.21 - 1.34 |
|  | October 7th | 4.17 | <0.001 | 3.97 - 4.38 |
|  | Relative National Stability | Ref |  |  |

Throughout the 19-year follow-up period, females were 33% more likely to purchase anxiolytics (HR 1.33, 95% CI: 1.32-1.35), while individuals who had a history of treatment with psychiatric drugs were had an overall 3.30-fold increased risk of first-time purchase of benzodiazepines. Compared to central Israel, residents of the North had a slight increased risk of purchasing anxiolytics, while residents of the Southern district experienced a 10% reduction in risk (HR 0.90, 95% CI: 0.88-0.91) with a further decreased risk for first time purchase for residents near the Gazan border (Gaza envelope), demonstrating an HR of 0.77 (95% CI: 0.74-0.79). Being Ultra-Orthodox Jewish slightly decreased one’s risk of anxiolytic treatment initiations (HR 0.97, 95% CI: 0.95-0.99) throughout the period while being of the Arab socio-religious sector much more so (HR 0.73, 95% CI: 0.71-0.75). Although both high and low socioeconomic statuses affected the risk for treatment initiation compared to a medium SES in a statistically significant manner, the impact was not clinically significant.

Periods of national armed conflicts significantly increased the risk for anti-anxiety treatment. The risk of anxiolytic treatment initiation during military operations was found to be 28% higher (HR 1.28, 95% CI: 1.21-1.34) compared to periods of relative national stability, the risk during the Second Lebanon War was 44% higher compared to periods of national security stability (HR 1.44 95% CI: 1.27-1.62) (**Table 2**, **Figure 3**). The October 7^th^ War was the most significant armed conflict leading to purchases of anti-anxiety medications throughout the 19-years follow-up, with individuals at 317% increased risk for treatment initiation following these events, compared to periods of relative national stability (HR 4.17, 95% CI: 3.97-4.38).

Examining the place of residence revealed distinct impacts related to October 7th, compared to other conflicts. Residents of northern Israel were at a higher risk during the Second Lebanon War (HR 1.39, 95% CI: 1.12-1.72), in contrast to other periods of national security instability (**Table S1**). During military operations, it was the residents of southern Israel who faced an increased risk for anxiolytic usage, with an HR of 1.18 (95% CI: 1.05-1.33). Conversely, the residential region did not significantly influence anti-anxiety treatment patterns following October 7th among residents of southern or northern Israel, compared to individuals living in central Israel. Notably, residing in the Gaza envelope did not result in a significantly different risk for first-time anxiolytic purchases compared to residents of the central region (**Table S1**).

In addition to having a baseline increased risk for anti-anxiety treatment initiation, women faced an additional elevated risk for anxiolytic treatment during periods of national security threats. The hazard ratio for the interaction between female sex and military operations was 1.26 (95% CI: 1.16-1.37), whereas the hazard ratio following the events of the recent war was 1.81 (95% CI: 1.67-1.97), indicating an 81% increased risk for anxiolytic treatment among women after October 7th (**Table S1**).

In order to further explore the effect of exposures on different sub-populations, we stratified the population by different socially distinct age groups (**Tables S2-S6**) and by socio-religious sectors (**Tables S7-S10**). Similarly to the main analysis, residents of the South were less likely to purchase anxiolytics at all ages throughout the follow-up period. Both October 7^th^ and military operations significantly increased the risk for first time anxiolytic purchases across age groups, while the Second Lebanon War impacted the 25-39-year olds and 40-59-year-olds (HR 1.54, 95%CI: 1.34-1.77 and 1.4, 1.13-1.74, respectively, **Table S2**). The former particularly in the North (HR 1.53, 95%CI: 1.18-1.98, **Table S4**).

Examining the different socio-religious sectors, results demonstrate that among the Arab population a higher SES increases the likelihood of treatment initiation by 27% (HR 1.27, 95%CI: 1.11-1.46), whereas among the general Jewish and Ultra-Orthodox Jewish population SES has much smaller or no significance (**Table S7**). Contrastingly to the general Jewish population (HR 1.31, 95% CI: 1.24-1.39) the Ultra-Orthodox Jewish and Arab sectors were not at significant higher risk for treatment initiation during military operations with HRs of 1.13 (95% CI: 0.93-1.36) and 0.72 (95% CI: 0.52-0.98), respectively. Similar outcome was seen for the Second Lebanon War, though the point estimate for the Arab population was borderline significant with an HR of 1.66 (95% CI 0.9-3.08). Following the October 7^th^ events, all sectors were at higher risk for treatment initiation with anti-anxiety medications, with HRs of 4.46,

2.89 and 1.61 for the general Jewish, Ultra-Orthodox Jew and Arab populations (**Table S7**). When examining the interaction of October 7^th^ with each of these socio-religious sectors (**Table S1**), the relevant HRs are 0.62 for the Ultra-Orthodox Jewish population (95%CI 0.52-0.72) and 0.35 (0.27-0.47) for the Arab population.

### *Secondary analyses (*renewed reinitiated treatment)

Results were overall similar when examining renewed treatment with anxiolytics (**Table 3, Figure S1**), with 276% increased risk for reinitiating anti-anxiety medications following October 7^th^ (HR 3.76, 95%CI: 3.55-3.98), and 42% and 25% increase during the Second Lebanon War and military operations. Interestingly, residents of the Gazan envelope were at a higher risk for repeated purchases of anxiolytics in the weeks following October 7^th^ (**Tables S11-S15**).

**Table 3.** Cox regression analysis with adjusted Hazard Ratios (HRs) for renewed purchase of anxiolytic medications (secondary analysis) between 2006-2024.

| Variable | Category | HR | P-value | 95% CI |
| --- | --- | --- | --- | --- |
| Age group, yr. | 25-39 | 1.04 | 0.003 | 1.01 - 1.07 |
|  | 40-59 | 1.18 | <0.001 | 1.15 - 1.21 |
|  | 60+ | 1.29 | <0.001 | 1.25 - 1.33 |
|  | 21-24 | Ref |  |  |
| Biological sex | Female | 1.21 | <0.001 | 1.20 - 1.23 |
|  | Male | Ref |  |  |
| District | North | 1.03 | <0.001 | 1.02 - 1.05 |
|  | South | 0.93 | <0.001 | 0.91 - 0.95 |
|  | Gaza envelope | 0.85 | <0.001 | 0.81 - 0.89 |
|  | Center | Ref |  |  |
| SES | Low | 1.00 | 0.97 | 0.98 - 1.02 |
|  | Medium | Ref |  |  |
|  | High | 1.04 | <0.001 | 1.03 - 1.06 |
| Socio-religious sector | Ultra-Orthodox Jew | 1.06 | 0.001 | 1.02 - 1.09 |
|  | Arab | 0.76 | <0.001 | 0.73 - 0.8 |
|  | General Jewish | Ref |  |  |
| Use of psychiatric drugs | Yes | 1.97 | <0.001 | 1.94 - 2.00 |
|  | No | Ref |  |  |
| Armed conflict | Second Lebanon War | 1.42 | <0.001 | 1.27 - 1.59 |
|  | Military operation | 1.25 | <0.001 | 1.19 - 1.32 |
|  | October 7th | 3.76 | <0.001 | 3.55 - 3.98 |
|  | Relative National Stability | Ref |  |  |

### Supplementary Analyses

The exposure negative controls analyses yielded no clinically significant increased risk for anxiolytic treatment during yearly shifts of dates of armed conflicts, whether first initiation or renewed treatment (**Tables S16-S17**).

## Discussion

This is the longest-spanning real-world study examining the effect of armed national conflicts in Israel on the mental health of the population, particularly focusing on stress- and anxiety-related reactions necessitating pharmaceutical intervention with short-term anxiolytics. We analyzed 19-years of medication purchase data of over 1.1 million members of a large-scale health organization.

Our analysis demonstrated that periods of national armed conflict significantly increased the risk for anti-anxiety treatment initiation. The risk for first purchase of an anxiety-relief short terms medication during military operations was found to be 28% higher (HR 1.28, 95% CI: 1.21-1.34) compared to periods of relative national stability, after adjusting for sociodemographic variables and previous psychiatric medical treatment. Higher risks were found following the two wars occurring during this time, with a 44% increased risk during the Second Lebanon War in 2006. The recent events of October 7^th^ were the most significant armed conflict increasing the risk for anxiety related reaction necessitating treatment throughout the 19-years follow-up, with individuals at 317% increased risk for treatment initiation compared to periods of relative national stability. The findings indicate that the recent war (October 7th) was the most significant variable influencing treatment initiation, demonstrating a greater impact than the effects observed for biological sex, age, previous psychiatric medication use, residential location, socioeconomic factors, and socio-religious affiliation. These results match some of the previous literature published, such as the systematic review of Ayer et al which found a correlation between exposure to armed conflicts and mental well-being. However, this study is not directly comparable, and other literature is limited, with a lack of longitudinal follow-up.^16^ This significant recent rise in anti-anxiety medication use, across different residential areas and population sectors, may suggest that the national anxiety level surpassed the capacity of traditional support systems, such as family, friends, and community networks, resulting in increased reliance on medications as a means of managing the immediate psychological impact. Contributing factors might include the pressure on national services during the crisis, challenges in emergency situation management, and the heightened stress experienced by healthcare professionals, who were also likely affected by the widespread distress.

Examining sociodemographic factors, data demonstrated that alongside a baseline increased risk for initiating anti-anxiety treatment, women experienced an additional elevated risk for anxiolytic therapy during times of national security threats, with 26% additional increased risk during military operations and an 81% increased risk for anxiolytic treatment among women after October 7th. Generally, older age groups were at a higher risk for anxiolytic treatment initiation, compared to the younger ones. Area of residence impacted treatment patterns. Overall, residents of northern Israel had a slight increased risk of purchasing anxiolytics compared to central Israel, while residents of the southern district had an 10% reduction in risk throughout the study period with a further decreased risk for first time purchase for residents near the Gazan border (Gaza envelope), demonstrating an HR of 0.77 (95% CI: 0.74-0.79).

During the Second Lebanon War, residents of northern Israel were at particularly higher risk for treatment initiation (HR 1.39, 95% CI: 1.12-1.72), unlike other conflicts, which might be explained by the fact that during the former, rockets were fired to civilian areas particularly in northern Israel, whereas in the latter, many of the involved civilian areas were in the South. Indeed, residents of southern Israel were at an increased risk during military operations (HR 1.18, 95% CI: 1.05-1.33). These finding somewhat align with previous studies, such as that of Gelhopf et al,^25^ demonstrated higher vulnerability to post-traumatic disorders in Sderot, a city in Southern Israel, though importantly, Gelhopf did not examine short-term effects.

Contrastingly, neither residents of southern Israel nor those near the Gaza border were at a significantly increased risk for anti-anxiety treatment initiation following October 7^th^ compared to residents of the central region. These findings somewhat depart from previous studies, such as the findings from Green et al’s systematic review of studies examining psychopathology of the southern adult population, exposed to continuous traumatic stress due to ongoing conflict. The reviewed studies reported high levels of probable posttraumatic stress disorder, depression, and other psychopathological reactions during low-intensity periods, which appear to rise sharply during escalations. However, these findings are somewhat limited by the lack of longitudinal data and the effect of changing intensity of exposure to the risk for developing anxiety and post traumatic mental health challanges.^10^ Moreover, our finding regarding the nationwide effect of October 7^th^ could be explained by the recent war’s overarching impact on the Israeli population, reflecting the violated sense of safety beyond residential district.

Interestingly, both the Ultra-Orthodox Jewish population and the Arab population had an overall significant decreased risk of initiating anxiolytics, especially pronounced among the latter population (HR 0.73, 95% CI: 0.71-0.75). This could be explained by social factors, such as stigma regarding seeking medical assistance in mental health challenges, or reduced access to community-level healthcare services. When stratifying by socio-religious sector, it is evident that higher SES plays a significant role among the Arab population, where a higher SES increases the likelihood of treatment initiation by 27%, whereas among the general Jewish and Ultra-Orthodox Jewish population SES has much smaller or no significance. Unlike the general Jewish population, the Ultra-Orthodox and Arab populations were not found to be at a significant higher risk for treatment initiation during military operations. Following the events of October 7^th^, however, all sectors were at higher risk for treatment initiation with anti-anxiety medications, with HRs of 4.46, 2.89 and 1.61 for the general Jewish, Ultra-Orthodox Jew and Arab populations respectively.

Examining renewed anti-anxiety treatment after at least one year washout period (of no treatment) yielded overall similar results to treatment initiation in terms of sociodemographic factors, with overall weaker associations to anxiolytic treatment. The events of October 7^th^ did increase the risk for renewed treatment, and more so in residents near the Gaza border, with an HR of the interaction term at 1.44 (95% CI: 1.08-1.94), which might be explained by that populations’ relapsing-remitting pattern of treatment and exposure to conflict.

Our analysis is subject to limitations. First, are analysis only included prescribed medications rather than off the counter (OTC) ones, which might provide further insight into individuals seeking medical intervention. However, MHS does not have access to OTC usage. Second, we opted to account for medication usage alone rather than other forms of treatment (such as mental health counseling), which yields a partial view of anxiety-related reactions. Unfortunately, due to lack of resources on a national level, scheduling appointments with mental health professionals often takes time, which will not allow us to evaluate short-term reactions, our main research question. Short-term anxiolytics can be prescribed by the primary care physician, a much more available resource to patients. As for refraining from usage of anxiety-related coded diagnoses, in Israel, the purchase of anxiety-related medications does not always correspond to an ‘anxiety-related coded diagnoses’, possibly due to physicians’ alignment with patients’ wishes to avoid societal stigma yielding from such diagnoses on medical forms, especially in more traditional populations. Third, women were more prevalent in our cohort, likely due to the inclusion and exclusion criteria, mandating participants to join prior to age 25 in order to allow as complete of a medical history as possible for distinction between first and renewed purchases. In Israel, men are often discharged from active duty in the late 20’s, rejoining one of the civilian state mandated health-funds. To mitigate this, analyses adjusted for biological sex, while interactions with sex were also included. Lastly, as in any retrospective observational study, estimates could be biased if confounders were not adequately addressed. To mitigate potential differences between groups, we adjusted for sociodemographic factors and prior psychiatric treatment. Moreover, we employed a Negative Controls approaches,^24^ yielding no clinically significant increased risk for anxiolytic treatment during yearly shifts of dates of armed conflicts, whether first initiation or renewed treatment, further strengthening the reliability of our conclusions. However, we recognize residual confounding could still be present, as in any retrospective study.

In conclusion, this study provides a comprehensive 19-year analysis of the relationship between national armed conflicts and short-term anxiolytic treatments in Israel. The findings reveal a significant increase in anxiety-related pharmacological intervention during periods of national security instability, most prominently after the recent events of October 7^th^. The heightened use of anxiolytics points to the substantial psychological impact of these conflicts, underscoring the need for targeted mental health interventions during such times. We must also consider that while anxiolytics could be effective for immediate relief, approaches focusing on long-term mental health and resilience are necessary to prevent dependency and improve mental health outcomes in the wake of these national crises.

## References

1. Javaid, S. F. et al. Epidemiology of anxiety disorders: global burden and sociodemographic associations. Middle East Current Psychiatry 30, 1–11 (2023).

2. Abrignani, M. G. et al. Panic disorder, anxiety, and cardiovascular diseases. Clin Neuropsychiatry 11, (2014).

3. Batelaan, N. M., ten Have, M., van Balkom, A. J. L. M., Tuithof, M. & de Graaf, R. Anxiety disorders and onset of cardiovascular disease: The differential impact of panic, phobias and worry. J Anxiety Disord 28, (2014).

4. Ryan, I. Anxious Generation: a Review of the Relationship Between Anxiety Disorders and Cardiovascular Disease in Youth. Curr Epidemiol Rep 7, (2020).

5. Pan, Y. et al. Association between anxiety and hypertension: A systematic review and meta-analysis of epidemiological studies. Neuropsychiatr Dis Treat 11, (2015).

6. Agbir, T., Okpara, I., Mbaave, P., Audu MD & Obindo, J. Generalised Anxiety Disorder and Cardiovascular Disease: A Study at a University Teaching Hospital in North-Central Nigeria. Journal of Research in Basic & Clinical Sciences | 1, (2019).

7. Gimson, A., Schlosser, M., Huntley, J. D. & Marchant, N. L. Support for midlife anxiety diagnosis as an independent risk factor for dementia: A systematic review. BMJ Open 8, (2018).

8. Gulpers, B. et al. Anxiety as a Predictor for Cognitive Decline and Dementia: A Systematic Review and Meta-Analysis. American Journal of Geriatric Psychiatry vol. 24 Preprint at 10.1016/j.jagp.2016.05.015 (2016).

9. Wittchen, H. U., Carter, R. M., Pfister, H., Montgomery, S. A. & Kessler, R. C. Disabilities and quality of life in pure and comorbid generalized anxiety disorder and major depression in a national survey. Int Clin Psychopharmacol 15, (2000).

10. Greene, T., Itzhaky, L., Bronstein, I. & Solomon, Z. Psychopathology, risk, and resilience under exposure to continuous traumatic stress: A systematic review of studies among adults living in Southern Israel. Traumatology (Tallahass Fla) 24, (2018).

11. Zvi, L. & Cohen-Louck, K. Exposure to continuous political violence: rational and experiential thinking styles, coping styles and post traumatic stress symptoms. Front Psychol 14, 1113608 (2023).

12. Besser, A., Weinberg, M., Zeigler-Hill, V. & Neria, Y. Acute symptoms of posttraumatic stress and dissociative experiences among female israeli civilians exposed to war: the roles of intrapersonal and interpersonal sources of resilience. J Clin Psychol 70, 1227– 1239 (2014).

13. Weinberg, M., Besser, A., Campeas, M., Shvil, E. & Neria, Y. Reactions of civilians exposed to terrorism and war trauma in Israel: The role of intra- and interpersonal factors. in Advances in psychology research, Vol. 94 1–53 (Nova Science Publishers, Hauppauge, NY, US, 2012).

14. Ron, P. PTSD, ASD, Secondary-Traumatization, and Death-Anxiety among Civilians and Professionals as Outcomes of On-Going Wars, Terror Attacks and Military Operations: An Integrative View. Psychology 10, (2019).

15. Shechory-Bitton, M. & Cohen-Louck, K. An Israeli Model for Predicting Fear of Terrorism Based on Community and Individual Factors. J Interpers Violence 35, (2020).

16. Ayer, L. et al. Psychological Aspects of the Israeli–Palestinian Conflict: A Systematic Review. Trauma, Violence, and Abuse vol. 18 Preprint at 10.1177/1524838015613774 (2017).

17. Israel Central Bureau of Statistics. Characterization and Classification of Geographical Units by the Socio-Economic Level of the Population 2015.. (2019).

18. Amer, M. Critical discourse analysis of war reporting in the international press: the case of the Gaza war of 2008–2009. Palgrave Communications 2017 3:1 3, 1–11 (2017).

19. Orenes, P. Operation Pillar of Defence and the 2013 Israeli Elections: Defensive or Provocative Intervention? J Terror Res 5, (2014).

20. Malka, V., Ariel, Y. & Avidar, R. Fighting, worrying and sharing: Operation ‘Protective Edge’ as the first WhatsApp war. 10.1177/17506352156116108, 329–344 (2015).

21. Yavetz, G. & Bronstein, J. Cities under fire: Crisis communication on home front versus frontline cities’ Facebook pages during operation ‘guardian of the walls’. Journal of Contingencies and Crisis Management 31, 421–430 (2023).

22. Hamieh, C. S. & Mac Ginty, R. A very political reconstruction: governance and reconstruction in Lebanon after the 2006 war. Disasters 34, S103–S123 (2010).

23. Barnea, A. Israeli Intelligence Was Caught Off Guard: The Hamas Attack on 7 October 2023—A Preliminary Analysis. International Journal of Intelligence and CounterIntelligence 1–27 (2024) doi:10.1080/08850607.2024.2315546.

24. Lipsitch, M., Tchetgen Tchetgen, E. & Cohen, T. Negative controls: a tool for detecting confounding and bias in observational studies. Epidemiology 21, 383–388 (2010).

25. Gelkopf, M., Berger, R., Bleich, A. & Silver, R. C. Protective factors and predictors of vulnerability to chronic stress: A comparative study of 4 communities after 7 years of continuous rocket fire. Soc Sci Med 74, (2012).

26. Yitshak-Sade, M., Mendelson, N., Novack, V., Codish, S. & Liberty, I. F. The association between an increase in glucose levels and armed conflict-related stress: A population-based study. Scientific Reports 2020 10:1 10, 1–6 (2020).

27. Soskolne, V., Dekel, R. & Vinker, S. Glycemic control of diabetes patients under continuous rocket attacks. Disaster Mil Med 2, (2016).

28. Rubinstein, A., Koffler, M., Villa, Y. & Graff, E. The Gulf War and Diabetes Mellitus. Diabetic Medicine 10, (1993).

