## Supplementary material for "Coping with Conflict: Short-Term Anxiolytic Medication Use Amidst National Stress in Israel"

**Figure S1.** Forest Plot of HRs and 95% confidence intervals, secondary analyses.

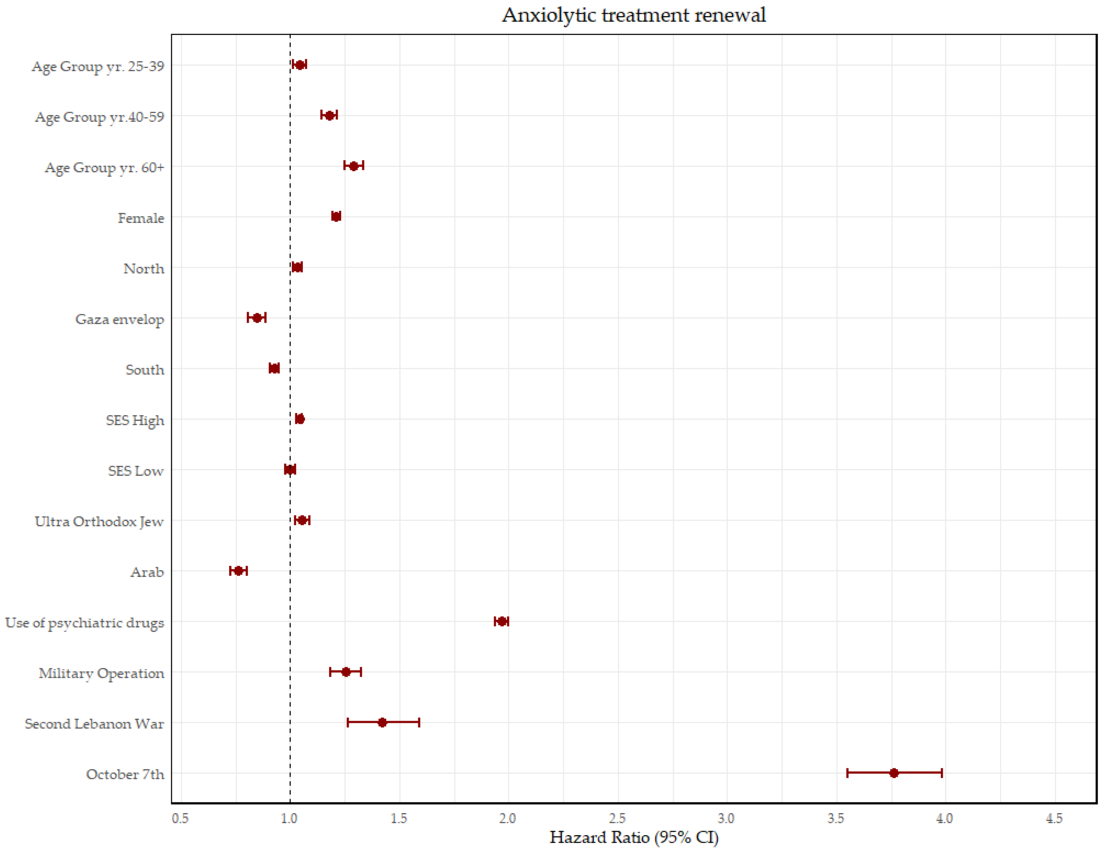


SES - socioeconomic status.

**Table S1.** Cox regression analysis with adjusted Hazard Ratios (HRs) for first purchase of anxiolytic medications (primary analysis) between 2006-2024.

| | Variable | Category | HR | P-value | 95% CI |
|---|---|---|---|---|---|
| Baseline | Age group, yr. | 25-39 | 1.25 | <0.001 | 1.22 - 1.29 |
| | | 40-59 | 1.68 | <0.001 | 1.63 - 1.72 |
| | | 60+ | 2.02 | <0.001 | 1.95 - 2.09 |
| | | 21-24 | Ref | | |
| | Biological sex | Female | 1.32 | <0.001 | 1.3 - 1.33 |
| | | Male | Ref | | |
| | District | North | 1.04 | <0.001 | 1.02 - 1.05 |
| | | South | 0.89 | <0.001 | 0.88 - 0.91 |
| | | Gaza envelope | 0.80 | <0.001 | 0.77 - 0.82 |
| | | Center | Ref | | |
| | SES | Low | 0.96 | <0.001 | 0.95 - 0.98 |
| | | Medium | Ref | | |
| | | High | 1.02 | 0.004 | 1.01 - 1.03 |
| | Socio-religious sector | Ultra-Orthodox Jew | 0.95 | <0.001 | 0.93 - 0.97 |
| | | Arab | 0.74 | <0.001 | 0.72 - 0.76 |
| | | General Jewish | Ref | | |
| | Use of psychiatric drugs | Yes | 3.29 | <0.001 | 3.26 - 3.33 |
| | | No | Ref | | |
| | Armed conflict | Second Lebanon War | 1.27 | 0.011 | 1.06 - 1.52 |
| | | Military operation | 1.14 | 0.002 | 1.05 - 1.23 |
| | | October 7th | 3.04 | <0.001 | 2.80 - 3.30 |
| Interaction with the Second Lebanon War | Biological sex | Female | 1.15 | 0.14 | 0.96 - 1.37 |
| | | Male | Ref | | |
| | District | North | 1.39 | 0.003 | 1.12 - 1.72 |
| | | South | 0.84 | 0.290 | 0.61 - 1.16 |
| | | Gaza envelope | 0.75 | 0.400 | 0.39 - 1.46 |
| | | Center | Ref | | |
| | Socio-religious sector | Ultra-Orthodox Jew | 0.92 | 0.634 | 0.55 - 1.18 |
| | | Arab | 1.05 | 0.822 | 0.54 - 1.45 |
| | | General Jewish | Ref | | |
| Interaction with Military operation | Biological sex | Female | 1.26 | <0.001 | 1.16 - 1.37 |
| | | Male | Ref | | |
| | District | North | 0.86 | 0.009 | 0.76 - 0.96 |
| | | South | 1.18 | 0.007 | 1.05 - 1.33 |
| | | Gaza envelope | 1.03 | 0.800 | 0.81 - 1.31 |
| | | Center | Ref | | |

| | | | | | |
|---|---|---|---|---|---|
| | Socio-religious sector | Ultra-Orthodox Jew | 0.83 | 0.023 | 0.71 - 0.98 |
| | | Arab | 0.59 | <0.001 | 0.45 - 0.79 |
| | | General Jewish | Ref | | |
| Interaction with October 7th | Biological sex | Female | 1.81 | <0.001 | 1.67 - 1.97 |
| | | Male | Ref | | |
| | District | North | 1.05 | 0.326 | 0.95 - 1.17 |
| | | South | 0.94 | 0.353 | 0.84 - 1.07 |
| | | Gaza envelope | 1.10 | 0.376 | 0.89 - 1.36 |
| | | Center | Ref | | |
| | Socio-religious sector | Ultra-Orthodox Jew | 0.62 | <0.001 | 0.52 - 0.72 |
| | | Arab | 0.36 | <0.001 | 0.27 - 0.47 |
| | | General Jewish | Ref | | |

HR – hazard ratio; CI – confidence interval; SES – socioeconomic status; yr – years.

**Table S2.** Cox regression analysis with adjusted Hazard Ratios (HRs) for first purchase of anxiolytic medications (primary analysis) between 2006-2024 – **stratified by age group.**

| Variable | Category | Age 21 - 24 | Age 25 - 39 | Age 40 - 59 | Age 60+ |
|---|---|---|---|---|---|
| Biological sex | Female | 1.36(1.33-1.4) | 1.28(1.27-1.3) | 1.39(1.37-1.42) | 1.36(1.3-1.42) |
| | Male | Ref | Ref | Ref | Ref |
| District | North | 1.13(1.09-1.17) | 1.01(1-1.03) | 1.02(1-1.05) | 1.01(0.94-1.08) |
| | South | 0.91(0.87-0.94) | 0.89(0.87-0.91) | 0.9(0.87-0.92) | 0.86(0.77-0.97) |
| | Gaza envelope | 0.71(0.66-0.77) | 0.82(0.78-0.85) | 0.81(0.76-0.86) | 0.79(0.59-1.07) |
| | Center | Ref | Ref | Ref | Ref |
| SES | Low | 0.99(0.95-1.03) | 0.96(0.94-0.98) | 0.95(0.92-0.98) | 1.01(0.91-1.11) |
| | Medium | Ref | Ref | Ref | Ref |
| | High | 1.09(1.06-1.13) | 1.01(0.99-1.02) | 1(0.98-1.02) | 1.03(0.98-1.07) |
| Socio-religious sector | Ultra-Orthodox Jew | 0.82(0.78-0.86) | 0.92(0.9-0.95) | 1.04(1-1.08) | 1.02(0.92-1.13) |
| | Arab | 0.61(0.57-0.66) | 0.76(0.74-0.8) | 0.76(0.72-0.8) | 0.76(0.63-0.92) |
| | General Jewish | Ref | Ref | Ref | Ref |
| Use of psychiatric drugs | Yes | 4.36(4.24-4.47) | 3.52(3.48-3.57) | 2.74(2.69-2.78) | 2.32(2.22-2.42) |
| | No | Ref | Ref | Ref | Ref |
| Armed conflict | Second Lebanon War | 1.03(0.75-1.42) | 1.54(1.34-1.77) | 1.4(1.13-1.74) | 1.11(0.72-1.71) |
| | Military operation | 1.32(1.19-1.47) | 1.28(1.21-1.36) | 1.27(1.18-1.37) | 1.44(1.21-1.71) |
| | October 7th | 3.32(2.99-3.68) | 4.55(4.3-4.81) | 3.91(3.64-4.2) | 2.67(2.1-3.4) |
| | Relative National Stability | Ref | Ref | Ref | Ref |

**Table S3.** Cox regression analysis with adjusted Hazard Ratios (HRs) for first purchase of anxiolytic medications (primary analysis) between 2006-2024, ages 21-24.

| | Variable | Category | HR | P-value | 95%CI |
|---|---|---|---|---|---|
| Baseline | Biological sex | Female | 1.35 | <0.001 | 1.32 - 1.39 |
| | | Male | Ref | | |
| | District | North | 1.13 | <0.001 | 1.09 - 1.17 |
| | | South | 0.90 | <0.001 | 0.87 - 0.94 |
| | | Gaza envelope | 0.71 | <0.001 | 0.66 - 0.77 |
| | | Center | Ref | | |
| | SES | Low | 0.99 | 0.588 | 0.95 - 1.03 |
| | | Medium | Ref | | |
| | | High | 1.09 | <0.001 | 1.06 - 1.13 |
| | Socio-religious sector | Ultra-Orthodox Jew | 0.83 | <0.001 | 0.78 - 0.87 |
| | | Arab | 0.62 | <0.001 | 0.58 - 0.67 |
| | | General Jewish | Ref | | |
| | Use of psychiatric drugs | Yes | 4.35 | <0.001 | 4.24 - 4.47 |
| | | No | Ref | | |
| | Armed conflict | Second Lebanon War | 0.89 | 0.683 | 0.49 - 1.59 |
| | | Military operation | 1.36 | 0.001 | 1.13 - 1.65 |
| | | October 7th | 2.63 | <0.001 | 2.17 - 3.21 |
| Interaction with the Second Lebanon War | Biological sex | Female | 1.03 | 0.918 | 0.55 - 1.95 |
| | | Male | Ref | | |
| | District | North | 1.54 | 0.247 | 0.74 - 3.21 |
| | | South | 0.90 | 0.844 | 0.31 - 2.57 |
| | | Gaza envelope | 0.00 | 0.942 | 0 - Inf |
| | | Center | Ref | | |
| | Socio-religious sector | Ultra-Orthodox Jew | 1.65 | 0.296 | 0.64 - 4.25 |
| | | Arab | 1.77 | 0.361 | 0.52 - 6.06 |
| | | General Jewish | Ref | | |
| Interaction with Military operation | Biological sex | Female | 1.06 | 0.588 | 0.86 - 1.32 |
| | | Male | Ref | | |
| | District | North | 0.78 | 0.118 | 0.58 - 1.06 |
| | | South | 1.05 | 0.784 | 0.76 - 1.44 |
| | | Gaza envelope | 0.87 | 0.672 | 0.46 - 1.65 |
| | | Center | Ref | | |
| | Socio-religious sector | Ultra-Orthodox Jew | 0.86 | 0.426 | 0.59 - 1.25 |
| | | Arab | 0.68 | 0.218 | 0.37 - 1.26 |
| | | General Jewish | Ref | | |
| Interaction with October 7th | Biological sex | Female | 1.54 | <0.001 | 1.24 - 1.9 |
| | | Male | Ref | | |

| | | | | |
|---|---|---|---|---|
| District | North | 1.02 | 0.869 | 0.79 - 1.33 |
| | South | 1.18 | 0.272 | 0.88 - 1.58 |
| | Gaza envelope | 1.37 | 0.224 | 0.82 - 2.29 |
| | Center | Ref | | |
| Socio-religious sector | Ultra-Orthodox Jew | 0.57 | 0.007 | 0.38 - 0.86 |
| | Arab | 0.32 | 0.003 | 0.15 - 0.68 |
| | General Jewish | Ref | | |

**Table S4.** Cox regression analysis with adjusted Hazard Ratios (HRs) for first purchase of anxiolytic medications (primary analysis) between 2006-2024, ages 25-39.

| | Variable | Category | HR | P-value | 95%CI |
|---|---|---|---|---|---|
| Baseline | Biological sex | Female | 1.27 | <0.001 | 1.25 - 1.28 |
| | | Male | Ref | | |
| | District | North | 1.01 | 0.194 | 0.99 - 1.03 |
| | | South | 0.89 | <0.001 | 0.87 - 0.91 |
| | | Gaza envelope | 0.82 | <0.001 | 0.78 - 0.85 |
| | | Center | Ref | | |
| | SES | Low | 0.96 | 0.001 | 0.94 - 0.99 |
| | | Medium | Ref | | |
| | | High | 1.01 | 0.53 | 0.99 - 1.02 |
| | Socio-religious sector | Ultra-Orthodox Jew | 0.93 | <0.001 | 0.9 - 0.96 |
| | | Arab | 0.78 | <0.001 | 0.75 - 0.81 |
| | | General Jewish | Ref | | |
| | Use of psychiatric drugs | Yes | 3.53 | <0.001 | 3.48 - 3.57 |
| | | No | Ref | | |
| | Armed conflict | Second Lebanon War | 1.3 | 0.017 | 1.05 - 1.62 |
| | | Military operation | 1.15 | 0.008 | 1.04 - 1.28 |
| | | October 7th | 3.22 | <0.001 | 2.89 - 3.58 |
| Interaction with the Second Lebanon War | Biological sex | Female | 1.17 | 0.174 | 0.94 - 1.45 |
| | | Male | Ref | | |
| | District | North | 1.53 | 0.001 | 1.18 - 1.98 |
| | | South | 0.85 | 0.402 | 0.59 - 1.24 |
| | | Gaza envelope | 0.97 | 0.932 | 0.5 - 1.89 |
| | | Center | Ref | | |
| | Socio-religious sector | Ultra-Orthodox Jew | 1.1 | 0.642 | 0.73 - 1.68 |
| | | Arab | 0.83 | 0.533 | 0.46 - 1.5 |
| | | General Jewish | Ref | | |
| Interaction with Military operation | Biological sex | Female | 1.26 | <0.001 | 1.13 - 1.42 |
| | | Male | Ref | | |
| | District | North | 0.85 | 0.055 | 0.72 - 1 |
| | | South | 1.15 | 0.092 | 0.98 - 1.36 |
| | | Gaza envelope | 1.13 | 0.416 | 0.84 - 1.52 |
| | | Center | Ref | | |
| | Socio-religious sector | Ultra-Orthodox Jew | 0.77 | 0.024 | 0.61 - 0.97 |
| | | Arab | 0.5 | 0.001 | 0.33 - 0.75 |
| | | General Jewish | Ref | | |

| | | | | | |
|---|---|---|---|---|---|
| Interaction with October 7th | Biological sex | Female | 1.87 | <0.001 | 1.67 - 2.1 |
| | | Male | Ref | | |
| | District | North | 1.12 | 0.134 | 0.97 - 1.29 |
| | | South | 0.86 | 0.091 | 0.72 - 1.03 |
| | | Gaza envelope | 1.02 | 0.905 | 0.76 - 1.36 |
| | | Center | Ref | | |
| | Socio-religious sector | Ultra-Orthodox Jew | 0.71 | 0.001 | 0.57 - 0.87 |
| | | Arab | 0.34 | <0.001 | 0.24 - 0.5 |
| | | General Jewish | Ref | | |

**Table S5.** Cox regression analysis with adjusted Hazard Ratios (HRs) for first purchase of anxiolytic medications (primary analysis) between 2006-2024, ages 40-59.

| | Variable | Category | HR | P-value | 95%CI |
|---|---|---|---|---|---|
| Baseline | Biological sex | Female | 1.37 | <0.001 | 1.35 - 1.4 |
| | | Male | Ref | | |
| | District | North | 1.03 | 0.042 | 1 - 1.05 |
| | | South | 0.89 | <0.001 | 0.87 - 0.92 |
| | | Gaza envelope | 0.81 | <0.001 | 0.77 - 0.86 |
| | | Center | Ref | | |
| | SES | Low | 0.95 | 0.001 | 0.92 - 0.98 |
| | | Medium | Ref | | |
| | | High | 1 | 0.958 | 0.98 - 1.02 |
| | Socio-religious sector | Ultra-Orthodox Jew | 1.05 | 0.017 | 1.01 - 1.09 |
| | | Arab | 0.77 | <0.001 | 0.73 - 0.82 |
| | | General Jewish | Ref | | |
| | Use of psychiatric drugs | Yes | 2.74 | <0.001 | 2.69 - 2.78 |
| | | No | Ref | | |
| | Armed conflict | Second Lebanon War | 1.36 | 0.104 | 0.94 - 1.96 |
| | | Military operation | 1.06 | 0.447 | 0.92 - 1.21 |
| | | October 7th | 2.74 | <0.001 | 2.38 - 3.16 |
| Interaction with the Second Lebanon War | Biological sex | Female | 1.12 | 0.58 | 0.75 - 1.67 |
| | | Male | Ref | | |
| | District | North | 1.21 | 0.468 | 0.72 - 2.03 |
| | | South | 0.86 | 0.745 | 0.35 - 2.12 |
| | | Gaza envelope | - | - | - |
| | | Center | Ref | | |
| | Socio-religious sector | Ultra-Orthodox Jew | 0.3 | 0.042 | 0.1 - 0.96 |
| | | Arab | 1.08 | 0.901 | 0.33 - 3.57 |
| | | General Jewish | Ref | | |
| Interaction with Military operation | Biological sex | Female | 1.33 | <0.001 | 1.14 - 1.55 |
| | | Male | Ref | | |
| | District | North | 0.91 | 0.384 | 0.74 - 1.12 |
| | | South | 1.32 | 0.013 | 1.06 - 1.63 |
| | | Gaza envelope | 0.86 | 0.58 | 0.52 - 1.45 |
| | | Center | Ref | | |
| | Socio-religious sector | Ultra-Orthodox Jew | 0.78 | 0.116 | 0.57 - 1.06 |
| | | Arab | 0.79 | 0.35 | 0.47 - 1.3 |
| | | General Jewish | Ref | | |
| Interaction with October 7th | Biological sex | Female | 1.83 | <0.001 | 1.57 - 2.13 |
| | | Male | Ref | | |

| | | | | |
|---|---|---|---|---|
| District | North | 0.95 | 0.592 | 0.79 - 1.14 |
| | South | 0.89 | 0.3 | 0.72 - 1.11 |
| | Gaza envelope | 1.06 | 0.774 | 0.72 - 1.56 |
| | Center | Ref | | |
| Socio-religious sector | Ultra-Orthodox Jew | 0.54 | 0.001 | 0.39 - 0.77 |
| | Arab | 0.35 | 0.001 | 0.19 - 0.64 |
| | General Jewish | Ref | | |

**Table S6.** Cox regression analysis with adjusted Hazard Ratios (HRs) for first purchase of anxiolytic medications (primary analysis) between 2006-2024, ages 60 or older.

| | Variable | Category | HR | P-value | 95%CI |
|---|---|---|---|---|---|
| Baseline | Biological sex | Female | 1.34 | <0.001 | 1.29 - 1.4 |
| | | Male | Ref | | |
| | District | North | 1.02 | 0.598 | 0.95 - 1.09 |
| | | South | 0.85 | 0.01 | 0.76 - 0.96 |
| | | Gaza envelope | 0.8 | 0.144 | 0.59 - 1.08 |
| | | Center | Ref | | |
| | SES | Low | 1.01 | 0.871 | 0.91 - 1.11 |
| | | Medium | Ref | | |
| | | High | 1.03 | 0.226 | 0.98 - 1.07 |
| | Socio-religious sector | Ultra-Orthodox Jew | 1.02 | 0.68 | 0.92 - 1.13 |
| | | Arab | 0.75 | 0.003 | 0.62 - 0.91 |
| | | General Jewish | Ref | | |
| | Use of psychiatric drugs | Yes | 2.32 | <0.001 | 2.22 - 2.42 |
| | | No | Ref | | |
| | Armed conflict | Second Lebanon War | 0.88 | 0.739 | 0.41 - 1.87 |
| | | Military operation | 1.06 | 0.733 | 0.76 - 1.48 |
| | | October 7th | 1.64 | 0.044 | 1.01 - 2.66 |
| Interaction with the Second Lebanon War | Biological sex | Female | 1.37 | 0.446 | 0.61 - 3.11 |
| | | Male | Ref | | |
| | District | North | 0.33 | 0.231 | 0.06 - 2.01 |
| | | South | 1.65 | 0.623 | 0.22 - 12.25 |
| | | Gaza envelope | - | - | - |
| | | Center | Ref | | |
| | Socio-religious sector | Ultra-Orthodox Jew | 0.92 | 0.907 | 0.22 - 3.88 |
| | | Arab | 16.34 | 0.002 | 2.68 - 99.56 |
| | | General Jewish | Ref | | |
| Interaction with Military operation | Biological sex | Female | 1.53 | 0.022 | 1.06 - 2.2 |
| | | Male | Ref | | |
| | District | North | 0.77 | 0.405 | 0.41 - 1.43 |
| | | South | 1.14 | 0.77 | 0.47 - 2.82 |
| | | Gaza envelope | 1.56 | 0.662 | 0.21 - 11.42 |
| | | Center | Ref | | |
| | Socio-religious sector | Ultra-Orthodox Jew | 1.45 | 0.136 | 0.89 - 2.36 |
| | | Arab | 0 | 0.948 | 0 - Inf |
| | | General Jewish | Ref | | |
| | Biological sex | Female | 2.05 | 0.006 | 1.23 - 3.42 |

| | | | | | |
|---|---|---|---|---|---|
| Interaction with October 7th | | Male | Ref | | |
| | District | North | 0.85 | 0.668 | 0.41 - 1.77 |
| | | South | 1.76 | 0.19 | 0.76 - 4.08 |
| | | Gaza envelope | - | - | - |
| | | Center | Ref | | |
| | Socio-religious sector | Ultra-Orthodox Jew | 0.4 | 0.117 | 0.13 - 1.26 |
| | | Arab | 2.01 | 0.283 | 0.56 - 7.22 |
| | | General Jewish | Ref | | |

**Table S7.** Cox regression analysis with adjusted Hazard Ratios (HRs) for first purchase of anxiolytic medications (primary analysis) between 2006-2024 – **stratified by socio-religious sector.**

| Variable | Category | General Jewish | Ultra-Orthodox Jewish | Arab |
|---|---|---|---|---|
| Age group, yr. | 25-39 | 1.22(1.19-1.25) | 1.51(1.38-1.65) | 1.47(1.3-1.67) |
| | 40-59 | 1.62(1.57-1.67) | 2.28(2.06-2.51) | 1.78(1.54-2.05) |
| | 60+ | 1.94(1.87-2.01) | 2.84(2.53-3.18) | 1.94(1.56-2.4) |
| | 21-24 | Ref | Ref | Ref |
| Biological sex | Female | 1.35(1.34-1.37) | 1.22(1.17-1.26) | 1.19(1.13-1.25) |
| | Male | Ref | Ref | Ref |
| District | North | 1.01(0.99-1.02) | 1.09(1.02-1.16) | 1.41(1.33-1.5) |
| | South | 0.79(0.77-0.82) | 0.76(0.67-0.86) | 2.02(0.65-6.29) |
| | Gaza envelope | 0.86(0.85-0.88) | 1.15(1.1-1.21) | 0.84(0.73-0.97) |
| | Center | Ref | Ref | Ref |
| SES | Low | 0.97(0.96-0.99) | 1.01(0.97-1.05) | 0.89(0.83-0.96) |
| | Medium | Ref | Ref | Ref |
| | High | 1.01(1-1.02) | 1.07(0.93-1.25) | 1.27(1.11-1.46) |
| Use of psychiatric drugs | Yes | 3.21(3.17-3.24) | 3.96(3.82-4.1) | 4.12(3.91-4.34) |
| | No | Ref | Ref | Ref |
| Armed conflict | Second Lebanon War | 1.45(1.27-1.65) | 1.19(0.77-1.83) | 1.66(0.9-3.08) |
| | Military operation | 1.31(1.24-1.39) | 1.13(0.93-1.36) | 0.72(0.52-0.98) |
| | October 7th | 4.46(4.23-4.7) | 2.89(2.39-3.49) | 1.61(1.15-2.27) |
| | Relative National Stability | Ref | Ref | Ref |

**Table S8.** Cox regression analysis with adjusted Hazard Ratios (HRs) for first purchase of anxiolytic medications (primary analysis) between 2006-2024, general Jewish population.

| | Variable | Category | HR | P-value | 95%CI |
|---|---|---|---|---|---|
| Baseline | Age group, yr. | 25-39 | 1.22 | <0.001 | 1.19 - 1.25 |
| | | 40-59 | 1.62 | <0.001 | 1.57 - 1.67 |
| | | 60+ | 1.94 | <0.001 | 1.87 - 2.01 |
| | | 21-24 | Ref | | |
| | Biological sex | Female | 1.34 | <0.001 | 1.32 - 1.35 |
| | | Male | Ref | | |
| | District | North | 1.01 | 0.404 | 0.99 - 1.02 |
| | | South | 0.86 | <0.001 | 0.85 - 0.88 |
| | | Gaza envelope | 0.79 | <0.001 | 0.77 - 0.81 |
| | | Center | Ref | | |
| | SES | Low | 0.97 | 0.006 | 0.96 - 0.99 |
| | | Medium | Ref | | |
| | | High | 1.01 | 0.045 | 1 - 1.02 |
| | Use of psychiatric drugs | Yes | 3.21 | <0.001 | 3.18 - 3.24 |
| | | No | Ref | | |
| | Armed conflict | Second Lebanon War | 1.29 | 0.008 | 1.07 - 1.57 |
| | | Military operation | 1.13 | 0.004 | 1.04 - 1.23 |
| | | October 7th | 2.98 | <0.001 | 2.74 - 3.24 |
| Interaction with the Second Lebanon War | Biological sex | Female | 1.12 | 0.23 | 0.93 - 1.36 |
| | | Male | Ref | | |
| | District | North | 1.35 | 0.01 | 1.07 - 1.7 |
| | | South | 0.88 | 0.444 | 0.62 - 1.23 |
| | | Gaza envelope | 0.79 | 0.493 | 0.41 - 1.54 |
| | | Center | Ref | | |
| Interaction with Military operation | Biological sex | Female | 1.26 | <0.001 | 1.15 - 1.37 |
| | | Male | Ref | | |
| | District | North | 0.87 | 0.023 | 0.77 - 0.98 |
| | | South | 1.18 | 0.012 | 1.04 - 1.34 |
| | | Gaza envelope | 1.04 | 0.74 | 0.82 - 1.33 |
| | | Center | Ref | | |
| Interaction with October 7th | Biological sex | Female | 1.82 | <0.001 | 1.67 - 1.99 |
| | | Male | Ref | | |
| | District | North | 1.08 | 0.168 | 0.97 - 1.2 |
| | | South | 0.95 | 0.445 | 0.84 - 1.08 |
| | | Gaza envelope | 1.12 | 0.287 | 0.91 - 1.39 |
| | | Center | Ref | | |

**Table S9.** Cox regression analysis with adjusted Hazard Ratios (HRs) for first purchase of anxiolytic medications (primary analysis) between 2006-2024, Ultra-Orthodox Jewish population.

| | Variable | Category | HR | P-value | 95%CI |
|---|---|---|---|---|---|
| Baseline | Age group, yr. | 25-39 | 1.51 | <0.001 | 1.38 - 1.65 |
| | | 40-59 | 2.28 | <0.001 | 2.07 - 2.51 |
| | | 60+ | 2.84 | <0.001 | 2.53 - 3.18 |
| | | 21-24 | Ref | | |
| | Biological sex | Female | 1.2 | <0.001 | 1.16 - 1.25 |
| | | Male | Ref | | |
| | District | North | 1.09 | 0.011 | 1.02 - 1.17 |
| | | South | 1.15 | <0.001 | 1.1 - 1.21 |
| | | Gaza envelope | 0.77 | <0.001 | 0.68 - 0.87 |
| | | Center | Ref | | |
| | SES | Low | 1.01 | 0.586 | 0.97 - 1.05 |
| | | Medium | Ref | | |
| | | High | 1.08 | 0.334 | 0.93 - 1.25 |
| | Use of psychiatric drugs | Yes | 3.96 | <0.001 | 3.82 - 4.1 |
| | | No | Ref | | |
| | Armed conflict | Second Lebanon War | 1.08 | 0.807 | 0.57 - 2.04 |
| | | Military operation | 0.96 | 0.773 | 0.73 - 1.27 |
| | | October 7th | 2.06 | <0.001 | 1.52 - 2.79 |
| Interaction with the Second Lebanon War | Biological sex | Female | 1.25 | 0.534 | 0.62 - 2.49 |
| | | Male | Ref | | |
| | District | North | 1.39 | 0.545 | 0.48 - 3.98 |
| | | South | 0.69 | 0.489 | 0.24 - 1.98 |
| | | Gaza envelope | 0 | 0.956 | 0 - Inf |
| | | Center | Ref | | |
| Interaction with Military operation | Biological sex | Female | 1.27 | 0.135 | 0.93 - 1.73 |
| | | Male | Ref | | |
| | District | North | 0.73 | 0.373 | 0.37 - 1.45 |
| | | South | 1.3 | 0.171 | 0.89 - 1.89 |
| | | Gaza envelope | 0.89 | 0.843 | 0.28 - 2.82 |
| | | Center | Ref | | |
| Interaction with October 7th | Biological sex | Female | 1.86 | <0.001 | 1.33 - 2.6 |
| | | Male | Ref | | |
| | District | North | 0.97 | 0.910 | 0.52 - 1.8 |
| | | South | 0.9 | 0.628 | 0.58 - 1.39 |
| | | Gaza envelope | 0.64 | 0.528 | 0.16 - 2.59 |
| | | Center | Ref | | |

**Table S10.** Cox regression analysis with adjusted Hazard Ratios (HRs) for first purchase of anxiolytic medications (primary analysis) between 2006-2024, Arab population.

| | Variable | Category | HR | P-value | 95%CI |
|---|---|---|---|---|---|
| Baseline | Age group, yr. | 25-39 | 1.47 | <0.001 | 1.3 - 1.67 |
| | | 40-59 | 1.78 | <0.001 | 1.54 - 2.05 |
| | | 60+ | 1.94 | <0.001 | 1.56 - 2.4 |
| | | 21-24 | Ref | | |
| | Biological sex | Female | 1.19 | <0.001 | 1.13 - 1.25 |
| | | Male | Ref | | |
| | District | North | 1.41 | <0.001 | 1.33 - 1.5 |
| | | South | 0.84 | 0.014 | 0.73 - 0.97 |
| | | Gaza envelope | - | - | - |
| | | Center | Ref | | |
| | SES | Low | 0.89 | 0.001 | 0.83 - 0.96 |
| | | Medium | Ref | | |
| | | High | 1.28 | <0.001 | 1.11 - 1.46 |
| | Use of psychiatric drugs | Yes | 4.12 | <0.001 | 3.91 - 4.34 |
| | | No | Ref | | |
| | Armed conflict | Second Lebanon War | 0.46 | 0.338 | 0.09 - 2.27 |
| | | Military operation | 0.68 | 0.214 | 0.37 - 1.25 |
| | | October 7th | 1.71 | 0.076 | 0.95 - 3.08 |
| Interaction with the Second Lebanon War | Biological sex | Female | 1.51 | 0.4 | 0.58 - 3.93 |
| | | Male | Ref | | |
| | District | North | 3.7 | 0.08 | 0.86 - 15.96 |
| | | South | 0 | 0.976 | 0 - Inf |
| | | Gaza envelope | - | - | - |
| | | Center | Ref | | |
| Interaction with Military operation | Biological sex | Female | 1.31 | 0.361 | 0.74 - 2.32 |
| | | Male | | | |
| | District | North | 0.84 | 0.566 | 0.47 - 1.51 |
| | | South | 0.95 | 0.946 | 0.22 - 4.13 |
| | | Gaza envelope | | 0.995 | |
| | | Center | Ref | | |
| Interaction with October 7th | Biological sex | Female | 1.031 | 0.916 | 0.59 - 1.81 |
| | | Male | Ref | | |
| | District | North | 0.84 | 0.566 | 0.47 - 1.52 |
| | | South | 1.98 | 0.275 | 0.58 - 6.8 |
| | | Gaza envelope | - | - | - |
| | | Center | Ref | | |

**Table S11.** Cox regression analysis with adjusted Hazard Ratios (HRs) for renewed purchase of anxiolytic medications (secondary analysis) between 2006-2024.

| | Variable | Category | HR | P-value | 95%CI |
|---|---|---|---|---|---|
| Baseline | Age group, yr. | 25-39 | 1.04 | 0.003 | 1.02 - 1.07 |
| | | 40-59 | 1.18 | <0.001 | 1.15 - 1.21 |
| | | 60+ | 1.29 | <0.001 | 1.25 - 1.33 |
| | | 21-24 | Ref | | |
| | Biological sex | Female | 1.20 | <0.001 | 1.18 - 1.21 |
| | | Male | Ref | | |
| | District | North | 1.03 | 0.004 | 1.01 - 1.05 |
| | | South | 0.92 | <0.001 | 0.9 - 0.95 |
| | | Gaza envelope | 0.85 | <0.001 | 0.81 - 0.89 |
| | | Center | Ref | | |
| | SES | Low | 0.99 | 0.251 | 0.97 - 1.01 |
| | | Medium | Ref | | |
| | | High | 1.04 | <0.001 | 1.03 - 1.06 |
| | Socio-religious sector | Ultra-Orthodox Jew | 1.08 | <0.001 | 1.05 - 1.11 |
| | | Arab | 0.83 | <0.001 | 0.79 - 0.86 |
| | | General Jewish | Ref | | |
| | Use of psychiatric drugs | Yes | 1.97 | <0.001 | 1.94 – 2.00 |
| | | No | Ref | | |
| | Armed conflict | Second Lebanon War | 1.23 | 0.054 | 1.0 - 1.52 |
| | | Military operation | 1.11 | 0.053 | 1.0 - 1.23 |
| | | October 7th | 2.42 | <0.001 | 2.16 - 2.73 |
| Interaction with the Second Lebanon War | Biological sex | Female | 1.21 | 0.100 | 0.97 - 1.51 |
| | | Male | Ref | | |
| | District | North | 1.28 | 0.079 | 0.97 - 1.69 |
| | | South | 0.74 | 0.187 | 0.48 - 1.16 |
| | | Gaza envelope | 0.73 | 0.483 | 0.3 - 1.77 |
| | | Center | Ref | | |
| | Socio-religious sector | Ultra-Orthodox Jew | 1.09 | 0.638 | 0.75 - 1.59 |
| | | Arab | 0.64 | 0.257 | 0.30 - 1.38 |
| | | General Jewish | Ref | | |
| Interaction with Military operation | Biological sex | Female | 1.18 | 0.004 | 1.05 - 1.32 |
| | | Male | Ref | | |
| | District | North | 1.07 | 0.393 | 0.92 - 1.24 |
| | | South | 1.17 | 0.082 | 0.98 - 1.39 |
| | | Gaza envelope | 1.04 | 0.837 | 0.73 - 1.49 |
| | | Center | Ref | | |

| | | | | | |
|---|---|---|---|---|---|
| | Socio-religious sector | Ultra-Orthodox Jew | 0.93 | 0.488 | 0.76 - 1.14 |
| | | Arab | 0.66 | 0.040 | 0.45 - 0.98 |
| | | General Jewish | Ref | | |
| Interaction with October 7th | Biological sex | Female | 1.89 | <0.001 | 1.67 - 2.13 |
| | | Male | Ref | | |
| | District | North | 1.15 | 0.062 | 0.99 - 1.32 |
| | | South | 1.06 | 0.483 | 0.9 - 1.26 |
| | | Gaza envelope | 1.44 | 0.015 | 1.08 - 1.94 |
| | | Center | Ref | | |
| | Socio-religious sector | Ultra-Orthodox Jew | 0.61 | <0.001 | 0.48 - 0.77 |
| | | Arab | 0.34 | <0.001 | 0.21 - 0.55 |
| | | General Jewish | Ref | | |

**Table S12.** Cox regression analysis with adjusted Hazard Ratios (HRs) for renewed purchases of anxiolytic medications (secondary analysis) between 2006-2024, ages 21-24.

| | Variable | Category | HR | P-value | 95%CI |
|---|---|---|---|---|---|
| Baseline | Biological sex | Female | 1.22 | <0.001 | 1.16 - 1.29 |
| | | Male | Ref | | |
| | District | North | 1.05 | 0.148 | 0.98 - 1.13 |
| | | South | 1.01 | 0.772 | 0.93 - 1.1 |
| | | Gaza envelope | 0.76 | 0.001 | 0.64 - 0.89 |
| | | Center | Ref | | |
| | SES | Low | 0.95 | 0.19 | 0.87 - 1.03 |
| | | Medium | Ref | | |
| | | High | 1.05 | 0.143 | 0.99 - 1.11 |
| | Socio-religious sector | Ultra-Orthodox Jew | 0.98 | 0.712 | 0.89 - 1.08 |
| | | Arab | 0.74 | <0.001 | 0.63 - 0.86 |
| | | General Jewish | Ref | | |
| | Use of psychiatric drugs | Yes | 2.61 | <0.001 | 2.43 - 2.8 |
| | | No | Ref | | |
| | Armed conflict | Second Lebanon War | 1.02 | 0.974 | 0.29 - 3.64 |
| | | Military operation | 0.81 | 0.374 | 0.5 - 1.3 |
| | | October 7th | 2.00 | 0.002 | 1.3 - 3.08 |
| Interaction with the Second Lebanon War | Biological sex | Female | 0.98 | 0.982 | 0.23 - 4.16 |
| | | Male | Ref | | |
| | District | North | 0.69 | 0.729 | 0.08 - 5.74 |
| | | South | 0.88 | 0.908 | 0.11 - 7.37 |
| | | Gaza envelope | - | - | - |
| | | Center | Ref | | |
| | Socio-religious sector | Ultra-Orthodox Jew | 1.11 | 0.92 | 0.13 - 9.27 |
| | | Arab | - | - | - |
| | | General Jewish | Ref | | |
| Interaction with Military operation | Biological sex | Female | 1.41 | 0.199 | 0.84 - 2.37 |
| | | Male | Ref | | |
| | District | North | 0.96 | 0.913 | 0.49 - 1.9 |
| | | South | 1.21 | 0.581 | 0.61 - 2.42 |
| | | Gaza envelope | - | - | - |
| | | Center | Ref | | |
| | Socio-religious sector | Ultra-Orthodox Jew | 1.15 | 0.723 | 0.54 - 2.42 |
| | | Arab | 0.92 | 0.913 | 0.22 - 3.96 |
| | | General Jewish | Ref | | |
| Interaction with October 7th | Biological sex | Female | 1.34 | 0.227 | 0.84 - 2.14 |
| | | Male | Ref | | |

| | | | | | |
|---|---|---|---|---|---|
| | District | North | 1.01 | 0.969 | 0.56 - 1.81 |
| | | South | 0.81 | 0.581 | 0.38 - 1.71 |
| | | Gaza envelope | 2.39 | 0.097 | 0.85 - 6.7 |
| | | Center | Ref | | |
| | Socio-religious sector | Ultra-Orthodox Jew | 0.62 | 0.3 | 0.25 - 1.54 |
| | | Arab | 0.65 | 0.55 | 0.15 - 2.7 |
| | | General Jewish | Ref | | |

**Table S13.** Cox regression analysis with adjusted Hazard Ratios (HRs) for renewed purchases of anxiolytic medications (secondary analysis) between 2006-2024, ages 25-39.

| | Variable | Category | HR | P-value | 95%CI |
|---|---|---|---|---|---|
| Baseline | Biological sex | Female | 1.13 | <0.001 | 1.11 - 1.15 |
| | | Male | Ref | | |
| | District | North | 1.03 | 0.088 | 1 - 1.06 |
| | | South | 0.91 | <0.001 | 0.88 - 0.95 |
| | | Gaza envelope | 0.86 | <0.001 | 0.8 - 0.91 |
| | | Center | Ref | | |
| | SES | Low | 0.98 | 0.243 | 0.95 - 1.01 |
| | | Medium | Ref | | |
| | | High | 1.06 | <0.001 | 1.03 - 1.08 |
| | Socio-religious sector | Ultra-Orthodox Jew | 1.06 | 0.012 | 1.01 - 1.1 |
| | | Arab | 0.88 | <0.001 | 0.83 - 0.93 |
| | | General Jewish | Ref | | |
| | Use of psychiatric drugs | Yes | 2.22 | <0.001 | 2.16 - 2.27 |
| | | No | Ref | | |
| | Armed conflict | Second Lebanon War | 1.08 | 0.629 | 0.79 - 1.48 |
| | | Military operation | 1.1 | 0.253 | 0.94 - 1.28 |
| | | October 7th | 2.39 | <0.001 | 2 - 2.87 |
| Interaction with the Second Lebanon War | Biological sex | Female | 1.2 | 0.3 | 0.85 - 1.68 |
| | | Male | Ref | | |
| | District | North | 1.74 | 0.004 | 1.19 - 2.54 |
| | | South | 0.59 | 0.134 | 0.3 - 1.17 |
| | | Gaza envelope | 0.99 | 0.987 | 0.36 - 2.7 |
| | | Center | Ref | | |
| | Socio-religious sector | Ultra-Orthodox Jew | 1.03 | 0.938 | 0.54 - 1.95 |
| | | Arab | 0.69 | 0.42 | 0.28 - 1.71 |
| | | General Jewish | Ref | | |
| Interaction with Military operation | Biological sex | Female | 1.2 | 0.047 | 1 - 1.43 |
| | | Male | Ref | | |
| | District | North | 0.98 | 0.85 | 0.77 - 1.24 |
| | | South | 1.16 | 0.257 | 0.9 - 1.5 |
| | | Gaza envelope | 0.85 | 0.548 | 0.5 - 1.45 |
| | | Center | Ref | | |
| | Socio-religious sector | Ultra-Orthodox Jew | 1.09 | 0.559 | 0.81 - 1.48 |
| | | Arab | 0.66 | 0.147 | 0.37 - 1.16 |
| | | General Jewish | Ref | | |
| | Biological sex | Female | 2.1 | <0.001 | 1.74 - 2.54 |

| | | | | | |
|---|---|---|---|---|---|
| Interaction with October 7th | | Male | Ref | | |
| | District | North | 1.28 | 0.026 | 1.03 - 1.58 |
| | | South | 0.97 | 0.837 | 0.74 - 1.28 |
| | | Gaza envelope | 1.27 | 0.309 | 0.8 - 2.02 |
| | | Center | Ref | | |
| | Socio-religious sector | Ultra-Orthodox Jew | 0.71 | 0.049 | 0.5 – 1.0 |
| | | Arab | 0.30 | 0.001 | 0.15 - 0.61 |
| | | General Jewish | Ref | | |

**Table S14.** Cox regression analysis with adjusted Hazard Ratios (HRs) for renewed purchases of anxiolytic medications (secondary analysis) between 2006-2024, ages 40-59.

| | Variable | Category | HR | P-value | 95%CI |
|---|---|---|---|---|---|
| Baseline | Biological sex | Female | 1.3 | <0.001 | 1.27 - 1.33 |
| | | Male | Ref | | |
| | District | North | 1.05 | 0.001 | 1.02 - 1.08 |
| | | South | 0.92 | <0.001 | 0.89 - 0.96 |
| | | Gaza envelope | 0.86 | <0.001 | 0.8 - 0.93 |
| | | Center | Ref | | |
| | SES | Low | 0.97 | 0.108 | 0.94 - 1.01 |
| | | Medium | Ref | | |
| | | High | 1.05 | <0.000 | 1.03 - 1.07 |
| | Socio-religious sector | Ultra-Orthodox Jew | 1.15 | <0.001 | 1.1 - 1.21 |
| | | Arab | 0.77 | <0.001 | 0.72 - 0.83 |
| | | General Jewish | Ref | | |
| | Use of psychiatric drugs | Yes | 1.84 | <0.001 | 1.8 - 1.88 |
| | | No | Ref | | |
| | Armed conflict | Second Lebanon War | 1.33 | 0.121 | 0.93 - 1.91 |
| | | Military operation | 1.13 | 0.152 | 0.96 - 1.33 |
| | | October 7th | 2.6 | <0.001 | 2.18 - 3.1 |
| Interaction with the Second Lebanon War | Biological sex | Female | 1.18 | 0.406 | 0.8 - 1.72 |
| | | Male | Ref | | |
| | District | North | 0.81 | 0.456 | 0.46 - 1.42 |
| | | South | 1.55 | 0.181 | 0.82 - 2.96 |
| | | Gaza envelope | 0.88 | 0.900 | 0.12 - 6.31 |
| | | Center | Ref | | |
| | Socio-religious sector | Ultra-Orthodox Jew | 1.21 | 0.48 | 0.71 - 2.07 |
| | | Arab | 0.96 | 0.959 | 0.23 - 4.05 |
| | | General Jewish | Ref | | |
| Interaction with Military operation | Biological sex | Female | 1.18 | 0.073 | 0.99 - 1.41 |
| | | Male | Ref | | |
| | District | North | 1.14 | 0.254 | 0.91 - 1.42 |
| | | South | 1.13 | 0.393 | 0.86 - 1.49 |
| | | Gaza envelope | 1.38 | 0.226 | 0.82 - 2.32 |
| | | Center | Ref | | |
| | Socio-religious sector | Ultra-Orthodox Jew | 0.84 | 0.314 | 0.59 - 1.19 |
| | | Arab | 0.72 | 0.294 | 0.39 - 1.33 |
| | | General Jewish | Ref | | |
| Interaction with October 7th | Biological sex | Female | 1.91 | <0.001 | 1.58 - 2.3 |
| | | Male | Ref | | |

| | | | | |
|---|---|---|---|---|
| District | North | 1.05 | 0.658 | 0.85 - 1.29 |
| | South | 1.09 | 0.471 | 0.86 - 1.39 |
| | Gaza envelope | 1.4 | 0.114 | 0.92 - 2.12 |
| | Center | Ref | | |
| Socio-religious sector | Ultra-Orthodox Jew | 0.5 | 0.001 | 0.33 - 0.75 |
| | Arab | 0.35 | 0.007 | 0.17 - 0.75 |
| | General Jewish | Ref | | |

**Table S15.** Cox regression analysis with adjusted Hazard Ratios (HRs) for renewed purchases of anxiolytic medications (secondary analysis) between 2006-2024, ages 60 or older.

| | Variable | Category | HR | P-value | 95%CI |
|---|---|---|---|---|---|
| Baseline | Biological sex | Female | 1.18 | <0.001 | 1.14 - 1.23 |
| | | Male | Ref | | |
| | District | North | 0.97 | 0.388 | 0.92 - 1.04 |
| | | South | 0.94 | 0.228 | 0.85 - 1.04 |
| | | Gaza envelope | 0.75 | 0.035 | 0.57 - 0.98 |
| | | Center | Ref | | |
| | SES | Low | 0.99 | 0.902 | 0.91 - 1.09 |
| | | Medium | Ref | | |
| | | High | 1.05 | 0.015 | 1.01 - 1.08 |
| | Socio-religious sector | Ultra-Orthodox Jew | 1.06 | 0.242 | 0.96 - 1.16 |
| | | Arab | 0.92 | 0.32 | 0.77 - 1.09 |
| | | General Jewish | Ref | | |
| | Use of psychiatric drugs | Yes | 1.45 | <0.001 | 1.39 - 1.52 |
| | | No | Ref | | |
| | Armed conflict | Second Lebanon War | 1.57 | 0.069 | 0.97 - 2.55 |
| | | Military operation | 1.3 | 0.058 | 0.99 - 1.71 |
| | | October 7th | 2.13 | <0.001 | 1.38 - 3.27 |
| Interaction with the Second Lebanon War | Biological sex | Female | 1.07 | 0.805 | 0.64 - 1.8 |
| | | Male | Ref | | |
| | District | North | 1.82 | 0.078 | 0.94 - 3.56 |
| | | South | 0.64 | 0.663 | 0.09 - 4.66 |
| | | Gaza envelope | - | - | - |
| | | Center | Ref | | |
| | Socio-religious sector | Ultra-Orthodox Jew | 0.9 | 0.841 | 0.33 - 2.48 |
| | | Arab | - | - | - |
| | | General Jewish | Ref | | |
| Interaction with Military operation | Biological sex | Female | 1.04 | 0.792 | 0.77 - 1.42 |
| | | Male | Ref | | |
| | District | North | 1.29 | 0.26 | 0.83 - 2.02 |
| | | South | 1.63 | 0.157 | 0.83 - 3.21 |
| | | Gaza envelope | 2.41 | 0.226 | 0.58 - 9.95 |
| | | Center | Ref | | |
| | Socio-religious sector | Ultra-Orthodox Jew | 0.69 | 0.241 | 0.38 - 1.28 |
| | | Arab | 0.35 | 0.3 | 0.05 - 2.56 |
| | | General Jewish | Ref | | |
| Interaction with October 7th | Biological sex | Female | 1.42 | 0.144 | 0.89 - 2.27 |
| | | Male | Ref | | |

| | | | | |
|---|---|---|---|---|
| District | North | 0.74 | 0.435 | 0.34 - 1.59 |
| | South | 1.07 | 0.891 | 0.39 - 2.94 |
| | Gaza envelope | - | - | - |
| | Center | Ref | | |
| Socio-religious sector | Ultra-Orthodox Jew | 0.74 | 0.483 | 0.32 - 1.7 |
| | Arab | - | - | - |
| | General Jewish | Ref | | |

**Table S16.** Exposure negative control analysis. Cox regression analysis with adjusted Hazard Ratios (HRs) for first purchases of anxiolytic medications during shifted dates of armed conflicts between 2006-2024.

| | Variable | Category | HR | P-value | 95%CI |
|---|---|---|---|---|---|
| Baseline | Age group, yr. | 25-39 | 1.23 | <0.001 | 1.2 - 1.26 |
| | | 40-59 | 1.62 | <0.001 | 1.58 - 1.67 |
| | | 60+ | 1.95 | <0.001 | 1.89 - 2.02 |
| | | 21-24 | Ref | | |
| | Biological sex | Female | 1.33 | <0.001 | 1.32 - 1.35 |
| | | Male | Ref | | |
| | District | North | 1.03 | <0.001 | 1.02 - 1.05 |
| | | South | 0.89 | <0.001 | 0.88 - 0.91 |
| | | Gaza envelope | 0.80 | <0.001 | 0.78 - 0.83 |
| | | Center | Ref | | |
| | SES | Low | 0.96 | <0.001 | 0.95 - 0.98 |
| | | Medium | Ref | | |
| | | High | 1.02 | 0.002 | 1.01 - 1.03 |
| | Socio-religious sector | Ultra-Orthodox Jew | 0.94 | <0.001 | 0.92 - 0.96 |
| | | Arab | 0.73 | <0.001 | 0.7 - 0.75 |
| | | General Jewish | Ref | | |
| | Use of psychiatric drugs | Yes | 3.30 | <0.001 | 3.26 - 3.33 |
| | | No | Ref | | |
| | Negative Control | Yes | 0.95 | 0.033 | 0.91 - 1 |
| | | No | Ref | | |
| Interaction with Negative Control Exposure (NCE) | Biological sex | Female | 0.99 | 0.579 | 0.94 - 1.04 |
| | | Male | Ref | | |
| | District | North | 1.05 | 0.203 | 0.98 - 1.12 |
| | | South | 1.00 | 0.967 | 0.92 - 1.09 |
| | | Gaza envelope | 0.87 | 0.092 | 0.74 - 1.02 |
| | | Center | Ref | | |
| | Socio-religious sector | Ultra-Orthodox Jew | 0.99 | 0.896 | 0.91 - 1.09 |
| | | Arab | 1.04 | 0.539 | 0.91 - 1.19 |
| | | General Jewish | Ref | | |

**Table S17.** Exposure negative control analysis. Cox regression analysis with adjusted Hazard Ratios (HRs) for renewed purchases of anxiolytic medications during shifted dates of armed conflicts between 2006-2024.

| | Variable | Category | HR | P-value | 95%CI |
|---|---|---|---|---|---|
| Baseline | Age group, yr. | 25-39 | 1.04 | 0.003 | 1.01 - 1.07 |
| | | 40-59 | 1.18 | 0.00 | 1.14 - 1.21 |
| | | 60+ | 1.29 | 0.00 | 1.25 - 1.33 |
| | | 21-24 | Ref | | |
| | Biological sex | Female | 1.21 | 0.00 | 1.19 - 1.23 |
| | | Male | Ref | | |
| | District | North | 1.04 | 0.00 | 1.02 - 1.05 |
| | | South | 0.93 | 0.00 | 0.9 - 0.95 |
| | | Gaza envelope | 0.85 | 0.00 | 0.81 - 0.89 |
| | | Center | Ref | | |
| | SES | Low | 0.99 | 0.260 | 0.97 - 1.01 |
| | | Medium | Ref | | |
| | | High | 1.04 | 0.00 | 1.03 - 1.06 |
| | Socio-religious sector | Ultra-Orthodox Jew | 1.07 | 0.00 | 1.04 - 1.1 |
| | | Arab | 0.82 | 0.00 | 0.78 - 0.85 |
| | | General Jewish | Ref | | |
| | Use of psychiatric drugs | Yes | 1.97 | 0.00 | 1.94 - 2 |
| | | No | Ref | | |
| | Negative Control | Yes | 0.98 | 0.406 | 0.92 - 1.04 |
| | | No | Ref | | |
| Interaction with Negative Control Exposure (NCE) | Biological sex | Female | 1.01 | 0.708 | 0.95 - 1.08 |
| | | Male | Ref | | |
| | District | North | 0.94 | 0.159 | 0.85 - 1.03 |
| | | South | 1.03 | 0.666 | 0.92 - 1.14 |
| | | Gaza envelope | 1.14 | 0.229 | 0.92 - 1.4 |
| | | Center | Ref | | |
| | Socio-religious sector | Ultra-Orthodox Jew | 1.03 | 0.684 | 0.91 - 1.16 |
| | | Arab | 0.92 | 0.460 | 0.75 - 1.14 |
| | | General Jewish | Ref | | |
